# FUS Mislocalization Rewires a Cortical Gene Network to Drive Cognitive and Behavioral Impairment in ALS

**DOI:** 10.1101/2025.06.16.25329673

**Authors:** Raphaelle Cassel, Félicie Lorenc, Aurélie Bombardier, Claudia De Tapia, Stéphane Dieterle, Cláudio Gouveia Roque, Christopher A. Jackson, Geoffrey Stuart-Lopez, Caroline Rouaux, Simon J. Guillot, Marie-Christine Birling, Pascal Kessler, Maurizio Grassano, Bryan Traynor, Adriano Chio, Raju Roy, James Shorter, Fergal M. Waldron, Jenna M. Gregory, Hemali Phatnani, Luc Dupuis, Salim Megat

**Affiliations:** Université de Strasbourg, Inserm, Strasbourg Translational Neuroscience and Psychiatry, UMR-S1329, Centre de Recherches en Biomédecine; Strasbourg, France; Center for Genomics of Neurodegenerative Disease, New York Genome Center, 101 Avenue of the Americas, New York, NY 10013, USA; PHENOMIN-Institut Clinique de la Souris, Université de Strasbourg, Illkirch, France; Université de Strasbourg, INSERM, UMS 38, Imaging Core Facility PIC-STRA, CRBS, Strasbourg, France; ALS Center ‘‘Rita Levi Montalcini’’ Department of Neuroscience, University of Turin, Turin, Italy; Neuromuscular Diseases Research Section, National Institute on Aging (NIA), National Institutes of Health (NIH), Bethesda, MD 20892, USA; Department of Neurology, Johns Hopkins University School of Medicine, Baltimore, MD 21287, USA; NINDS, NIH, Bethesda, MD 20892, USA; ASO Therapeutics Laboratory, National Center for Advancing Translational Sciences, NIH, Rockville, MD 20850, USA; Department of Biochemistry and Biophysics, Perelman School of Medicine, University of Pennsylvania, Philadelphia, PA 19104. U.S.A; Institute of Medical Sciences, University of Aberdeen, UK; Department of Neurology, Center for Translational and Computational Neuroimmunology, Columbia University, New York, NY 10032, USA

**Author notes:** Equal contribution (Luc Dupuis, Salim Megat). Lead contact: Luc DUPUIS.

**Keywords:** Amyotrophic lateral sclerosis, fronto-temporal dementia, cognitive impairment, behavioral impairment, mouse models, genetics, single cell biology

## Abstract

Cognitive and behavioral impairment affects up to half of individuals with amyotrophic lateral sclerosis (ALS), but their molecular origin remains unresolved. Here, we identify mislocalization of the RNA-binding protein FUS in cortical neurons as a defining feature in ALS patients with cognitive impairment (ALS-ci). Selective mislocalization of FUS in adult cortical projection neurons in mice is sufficient to trigger ALS-ci– and ALS with behavioral impairment (ALS-bi)–like phenotypes, including deficits in sociability, and neurodegeneration. Single-nucleus transcriptomics reveal a conserved FUS-dependent gene network downregulated in these mice and ALS-ci patients. This regulon is enriched for ALS genetic risk factors and newly implicates *FBXO16* in ALS-bi. Carriers of protein-truncating *FBXO16* variants display behavioral abnormalities, frontotemporal atrophy, and increased levels of dementia-linked biomarkers. These findings define a neuron-intrinsic mechanism for cognitive and behavioral dysfunction in ALS and nominate FUS mislocalization and its downstream gene network as therapeutic targets.

**Graphical abstract:** 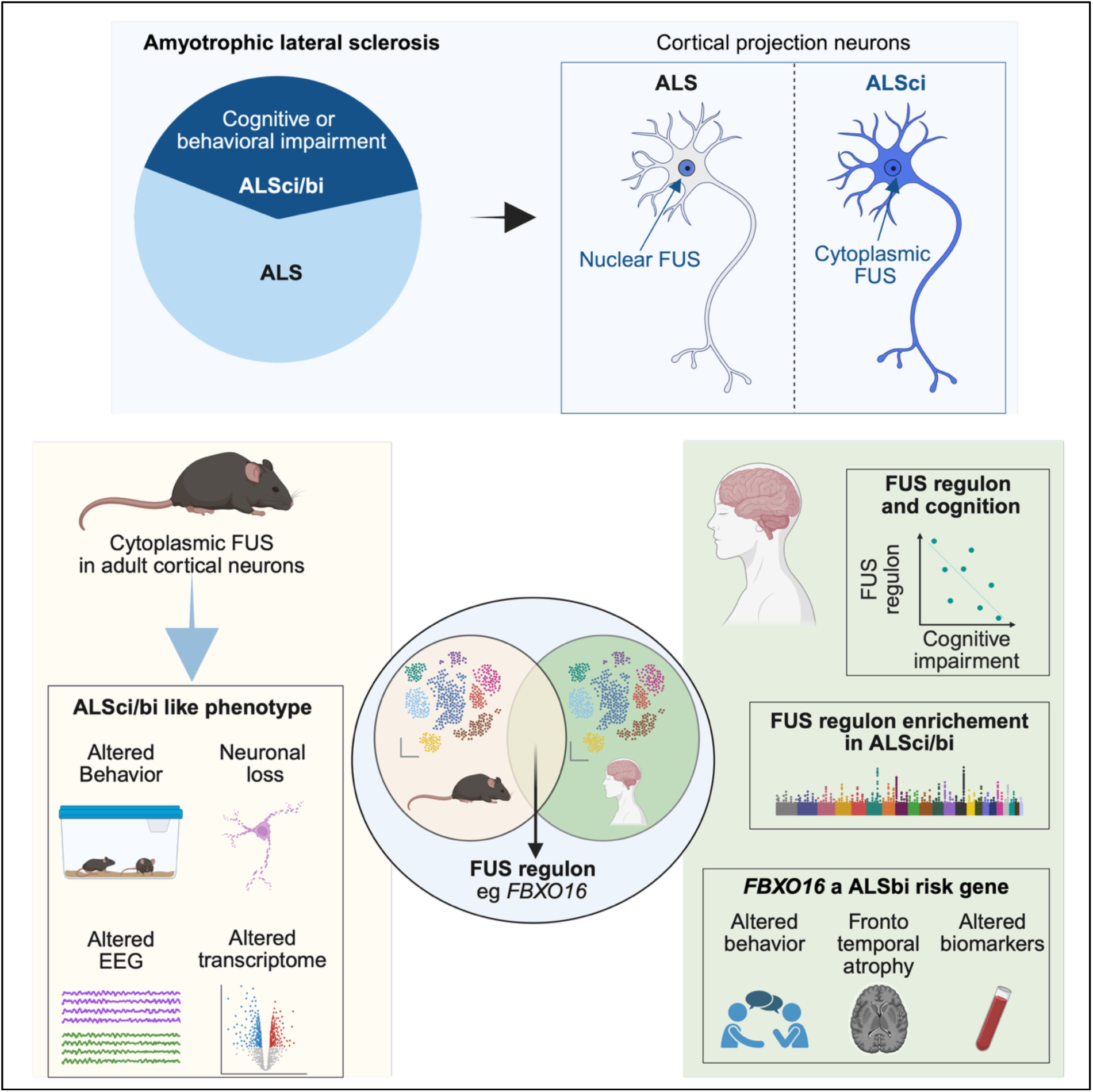

**Highlights:**

- Transcriptional fingerprint of FUS mislocalization is observed in cortical projection neurons of ALS patients
- FUS mislocalization leads to downregulation in cortical projection neurons of a cross-species conserved regulon shared with TDP43
- FUS mislocalization in adult cortical projection neurons is sufficient to trigger ALS related cognitive and behavioral impairment in mouse models
- FUS is mislocalized and the FUS regulon is downregulated in ALS patients with cognitive impairment
- The FUS regulon is enriched in genetic risk factors for cognitive and behavioral impairment in ALS
- Carriers of protein truncating variants of *FBXO16*, one of the FUS regulon genes, display behavioral, imaging and biochemical markers of ALS with behavioral impairment.

## Introduction

Amyotrophic lateral sclerosis (ALS) is the most frequent adult-onset disease of motor neurons. ALS is characterized by the onset of motor symptoms in the 5^th^ or 6^th^ decade of life, rapidly progressing to fatal paralysis, usually within two to three years after symptoms’ onset. A substantial fraction of ALS patients (up to 50%) develop additional symptoms, unrelated to the motor system, in particular variable cognitive impairment (ALS-ci) or behavioral impairment (ALS-bi), with some patients even presenting with frontotemporal dementia (ALS-FTD) (Abrahams, 2023). The relevance of this ALS-FTD clinical continuum is substantiated by the existence of a common pathology between ALS and FTD, in particular aggregates of TDP-43, an RNA-binding protein (RBP), as well as common genetic associations (Goutman et al., 2022a, b). It remains unclear, however, whether ALS-ci, ALS-bi and ALS without cognitive impairment have distinct mechanisms. Indeed, TDP-43 pathology in areas involved in cognition, behavior and executive functions is a poor predictor of ALS-ci or ALS-bi (Bampton et al., 2024; Rifai et al., 2024; Suzuki et al., 2024), suggesting that additional pathways contribute to the development of these extra-motor deficits.

An attractive hypothesis would be that other, yet undetermined, pathological features develop in ALS-ci or ALS-bi patients but not in patients without cognitive or behavioral impairment, to cause the ci/bi phenotypes. As a matter of fact, the co-occurrence of several proteinopathies is frequent in neurodegenerative diseases, and is generally associated with worsened prognosis (Irwin et al., 2017; James et al., 2016; Karanth et al., 2020; Katsumata et al., 2020; Robinson et al., 2023). Interestingly, the heritability of ALS is enriched in TDP-43 targets (Brown et al., 2022; Ling et al., 2015; Ma et al., 2022; Megat et al., 2023). Beyond TDP-43, the heritability of ALS is also enriched in targets of another RBP linked to ALS, FUS (Megat et al., 2023). Mutations in the *FUS* gene cause severe familial ALS cases through altered nuclear import of the FUS protein (Kwiatkowski et al., 2009; Moens et al., 2025; Scekic-Zahirovic et al., 2017; Scekic-Zahirovic et al., 2016; Vance et al., 2009). FUS aggregates are also observed in a subset of FTD cases, in complex aggregates that also include TAF15 and other RBPs (Mackenzie et al., 2011a; Mackenzie et al., 2011b; Neumann et al., 2011; Urwin et al., 2010). Importantly, FUS is also mislocalized in the cytoplasm of neurons in sporadic ALS, even in the absence of obvious FUS aggregates (Tyzack et al., 2019; Tyzack et al., 2021), as well as in *C9ORF72* ALS (Rifai et al., 2022). Despite these observations, the distribution of FUS mislocalization across ALS, ALS-ci and ALS-bi patients remains unknown, and the consequences of FUS mislocalization on ALS clinical features, including cognitive and behavioral, are poorly characterized.

Here, we demonstrate that FUS mislocalization in cortical neurons is more pronounced in ALS-ci than in ALS patients. Furthermore, FUS mislocalization in adult projection neurons is sufficient to lead to ALS-ci/bi-like phenotypes in mice. Through integration of various single-cell datasets, we identify a gene module downstream of FUS mislocalization that is downregulated in cortical neurons of ALS-ci patients and identify genetic associations between *FBXO16* and *WWOX,* two genes of this module, with ALS-bi and ALS-ci respectively. Thus, FUS mislocalization in projection neurons compromises expression of a gene network underlying the risk of cognitive and behavioral impairment in ALS.

## Results

### Mislocalization of FUS is sufficient to recapitulate transcriptomic alterations in ALS excitatory projection neurons

Previous studies have observed mislocalization of FUS in neurons of sporadic and familial ALS patients, including *C9ORF72* (Rifai et al., 2022; Tyzack et al., 2019; Tyzack et al., 2021), but did not define downstream transcriptional consequences. To identify those alterations, we generated a single-nuclei RNA sequencing (snRNAseq) dataset of the frontal cortex of *Fus*^ΔNLS/+^ mice. This mouse model is a knock-in mouse model expressing a FUS protein truncated of its C-terminal nuclear localization sequence (NLS). This leads to the cytoplasmic accumulation of FUS (Scekic-Zahirovic et al., 2017; Scekic-Zahirovic et al., 2016), without aggregation. Similar mutations have been documented in ALS patients, and the cytoplasmic accumulation of FUS without aggregation is similar to what has been reported in ALS patients (Rifai et al., 2022; Tyzack et al., 2019; Tyzack et al., 2021). We used this novel dataset as a blueprint of FUS mislocalization and compared it with a publicly available dataset of motor cortex of ALS patients (Pineda et al., 2024) (**Figure 1A-B**). Single nuclei RNA sequencing of frontal cortex of *Fus*^ΔNLS/+^ mice yielded 78,514 nuclei allowing the identification of 21 clusters that reflect large cellular subtypes as previously described in the mouse primary motor cortex (Network, 2021). Analysis of human snRNAseq from the motor cortex yielded 156,545 nuclei split across 19 subclasses that allow the identification of the major cellular subtypes such as upper and deep layer excitatory neurons, interneurons derived from the medial and caudal ganglionic eminences, as well as different types of non-neuronal cells such as microglia, astrocytes and endothelial cells (**Figure 1C-D**). Principal component analysis of *Fus*^ΔNLS/+^ mice and ALS patients showed strong transcriptional differences among excitatory projection neurons that is confirmed with a higher number of differentially expressed (DE) genes compared to inhibitory neurons and non-neuronal cells (**Figure 1C-D**). Gene ontology analysis performed on the overlapped DE genes in excitatory neurons in *Fus*^ΔNLS/+^ mice and ALS patients show a similar enrichment in terms associated with mitochondrial function, RNA binding and mRNA translation (FDR<0.05) suggesting a common underlying mechanism driving neurodegeneration in human ALS patient and *Fus*^ΔNLS/+^ mice. These results show that endogenous mislocalization of FUS leads to similar transcriptional alterations as ALS specifically in cortical projection neurons (CPNs) (**Figure 1E-F**).

**Figure 1:**
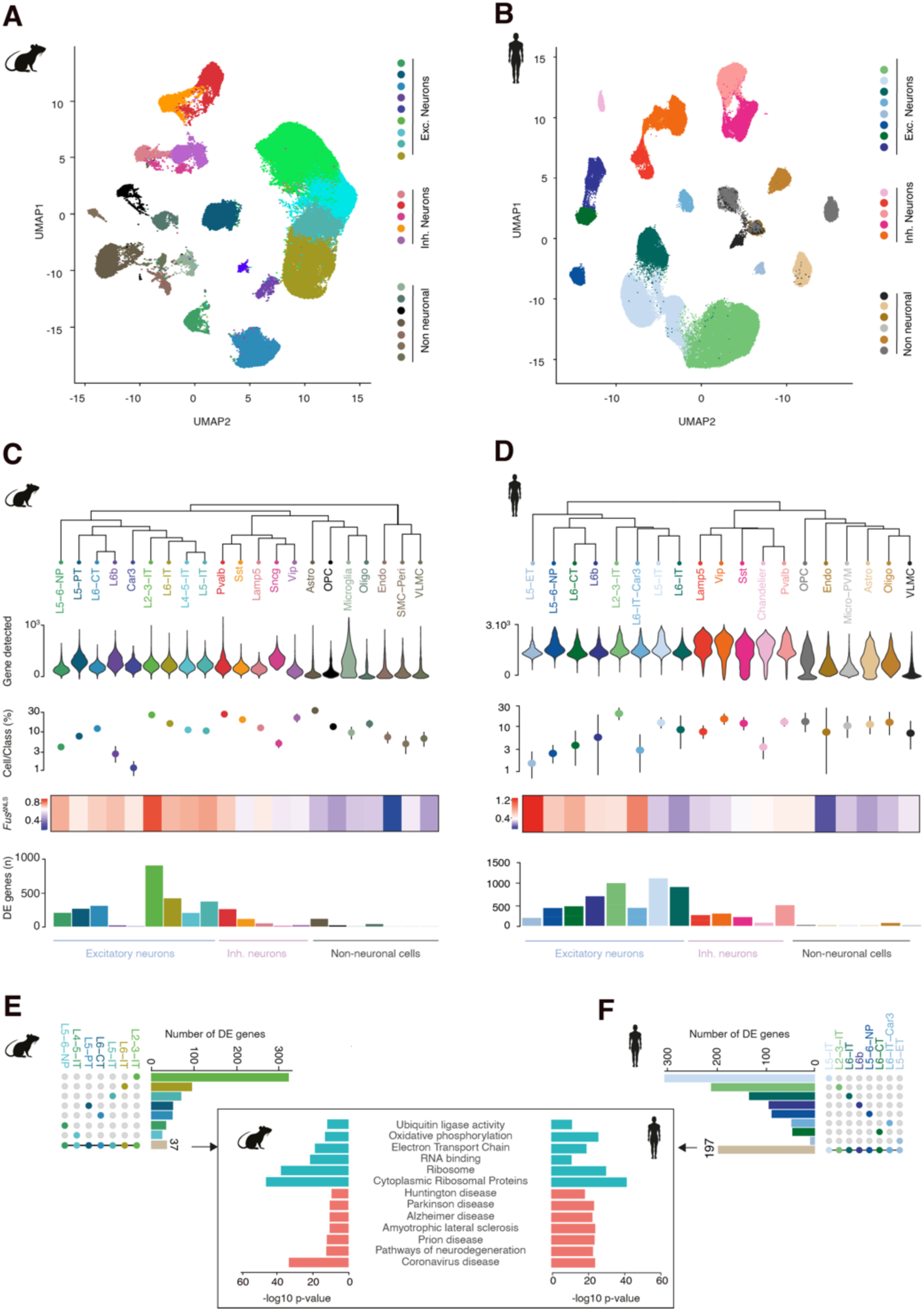
Cross-species transcriptomic analysis identifies FUS-related signature in vulnerable ALS neurons. **A.** UMAP plot of the 78,514 cells split across 21 clusters identified in the frontal cortex of Fus^ΔNLS/+^ mice **B.** UMAP plot of the 156,545 cells split across 19 clusters in the human M1 motor cortex. **C.** Cluster dendrogram, violin plots heatmaps and histogram plot shows number of genes detected, percent of each class (Glutamategic, GABAergic and non-neurons), transcriptome-wide shift from controls in Fus^ΔNLS/+^ mice and number of DE genes in each cell subclass. **D.** Cluster dendrogram, violin plots, heatmap and histogram bar plot shows number of genes detected, percent of each class (Glutamategic, GABAergic and non-neurons), transcriptome-wide shift of ALS patients compared to healthy donors and number of DE genes in each cell subclass. **E-F:** Intersect plots show the number of DE genes in each excitatory subclass and overlap used to perform gene ontology analysis in mice and human. Histogram bar plot shows gene ontology terms enriched in all excitatory neurons of ALS patients and Fus^ΔNLS/+^ mice.

### A conserved FUS regulon shared with TDP-43 in ALS-vulnerable neurons

FUS is an RNA binding protein with a prion-like domain and thousands of RNA targets (Lagier-Tourenne et al., 2012; Rogelj et al., 2012). To narrow down to the critical RNA targets associated with ALS, we reasoned that the genes of interest should (*i)* be modulated by FUS in CPNs and (*ii*) show similar alterations in mouse models and ALS patients. To integrate gene expression changes in a systems-level framework, we performed a co-expression analysis across available mouse and human datasets to identify significant co-expressed modules, using single-cell transcriptome of human L5 extra-telencephalic (L5-ET) vulnerable neurons (Pineda et al., 2024) and single-nuclei transcriptome of *Fus*^ΔNLS/+^ mice (**Figure 2A-B**). We identified a total of 4 gene modules with signed correlation to ALS in either L5-ET human neurons, or *Fus*^ΔNLS/+^ transcriptome, or both. Only one module, color coded “Turquoise”, showed correlation to ALS in both mouse and human species (**Figure 2B-E**). Specifically, the “Turquoise” module was downregulated in both human and mouse datasets (FDR <0.05) (**Figure 2E**). As we assume the module to be regulated through a FUS-dependent mechanism, we decided to name this module “FUS regulon” (**Table S1**). Gene ontology of the FUS regulon showed enrichment in genes involved in functions altered in ALS, e.g. cilium organization (De Decker et al., 2025), cytoskeleton or RNA binding (Brown and Al-Chalabi, 2017) (FDR<0.05) (**Figure 2F**). Downregulation of the FUS regulon was widespread across CPN in *Fus*^ΔNLS/+^ mice as shown in **Figure 2G** using eigengene of the FUS regulon as a global measurement of FUS regulon expression. For instance, the FUS regulon was downregulated in L5-PT neurons (grouping most extra-telencephalic neurons in mice) (unadjusted-p = 0.040), but also in L5-IT (intra-telencephalic neurons) (*Bonferroni corrected-p <0.05) in *Fus*^ΔNLS/+^ mice (**Figure 2G**). Downregulation of the FUS regulon was significant in L5-ET neurons of the motor cortex of sporadic ALS patients (*Bonferroni corrected-p <0.05) (**Figure 2H**). To determine the specificity of the FUS regulon towards FUS among other RBPs, we evaluated expression of the FUS regulon in a publicly available dataset (Van Nostrand et al., 2020) that explored transcriptional effects of downregulation of 217 RBPs (**Figure 2I**). In this dataset, FUS ranked 8^th^ in RBPs downregulating the FUS regulon (**Table S2**). Interestingly, TDP-43 ranked 2^nd^ as RBP with the most downregulation of the FUS regulon, suggesting that FUS and TDP-43 are both necessary for strong expression of the FUS regulon. Consistent with this, knockdown of TDP-43 in iPSC-derived motor neurons (Brown et al., 2022) led to decreased expression of the FUS regulon (**p<0.01) (**Figure 2J**), and expression of the FUS regulon was decreased in TDP-43 negative neuronal nuclei as compared to TDP-43 positive nuclei in FTD-TDP43 patients samples (***p<0.001) (Liu et al., 2019) (**Figure 2K**). These results demonstrate that FUS mislocalization leads to downregulation of the FUS regulon in CPNs, phenocopying transcriptional alterations in vulnerable neurons in patients and is associated with FUS and TDP43 loss of function.

**Figure 2:**
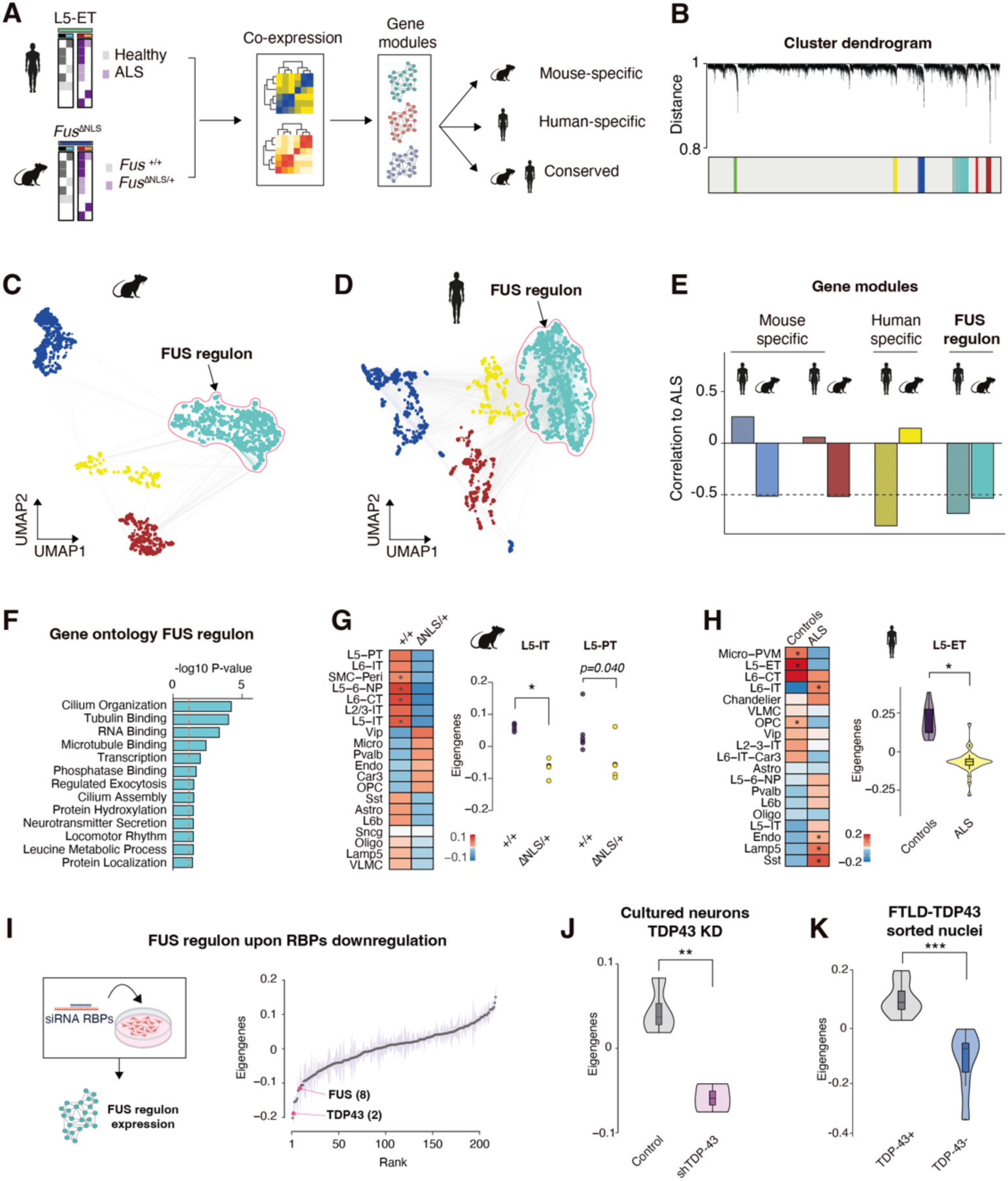
Cell-type specific gene module analysis identifies conserved regulatory networks associated with FUS, and shared with TDP-43. **A.** Experimental design of the gene network analysis in Fus^ΔNLS/+^ mice and ALS patients. **B.** Cluster dendrogam showing the gene modules identified in human ALS patients and Fus^ΔNLS/+^ mice. **C-D.** UMAP gene network of the selected modules, each dot represents individual gene which size is proportional to its importance (signed kME). The FUS regulon displays a high degree of co-expression in mice which is conserved in ALS patients. **E.** Bar graphs showing correlation of each module to ALS phenotype and directionality in mice and human. Dashed lines highlight correlation with FDR<0.05 (Benjamini-Hochberg correction). **F.** Histogram bar graphs show GO enrichment of the FUS regulon. Dashed lines highlight GO term with FDR<0.05 (Benjamini-Hochberg correction). **G.** Heatmap shows eigengene of the FUS regulon across all cell types in *Fus*^ΔNLS/+^ mice. Wilcoxon rank-sum test with Bonferroni correction *p< 0.05/19 (number of cell types). Dot plot shows FUS regulon eigengene in L5-IT neurons of *Fus*^ΔNLS/+^ mice (Wilcoxon rank-sum test with Bonferroni correction *p=0.00024) **H.** Heatmap shows FUS regulon eigengene across all cell types in ALS patients. Wilcoxon rank-sum test with Bonferroni correction *p< 0.05/21(number of cell types). Dot plot shows eigengenes expression in L5-PT neurons of Fus^ΔNLS/+^ mice (Wilcoxon rank-sum test with Bonferroni correction p=0.040) **I.** Eigengenes expression of the FUS regulon upon downregulation of 217 different RNA-binding proteins and ranked according to effect sizes. **J.** Violin plot showing FUS regulon eigengene expression in iPSC-derived motoneurons treated with TDP43 shRNA (one-way ANOVA: F_group_=31.39, **p=0.0025). **K.** Violin plot showing FUS regulon eigengene expression in TDP43^−^ nuclei from FTLD patients (one-way ANOVA: F_group_=21.14, ***p=0.000613).

### Adult-onset FUS mislocalization in mouse cortical projection neurons reproduces molecular, pathological and electrical alterations of ALS patients

As the transcriptional consequences of FUS mislocalization were most prominent in CPNs, we then sought to restrict FUS mislocalization in adult excitatory projection neurons in a mouse model. Thus, we generated a novel mouse *Fus* allele allowing conditional ablation of the nuclear localization signal (NLS), leading to cytoplasmic accumulation of FUS. We engineered the *Fus* locus to insert loxP sequences in intron 12 and downstream of exon 15 (**Figure S1A**). The second loxP site is followed by a cDNA encoding exons 13 and 14 of *Fus*, followed by 3 transcriptional stop cassettes and a poly-adenylation signal to restore exons 13 and 14 after CRE recombination (**Figure S1A**). Thus, CRE-mediated recombination is expected to delete exon 15 leading to a FUS protein without NLS, and thus to the same mutant FUS protein as in constitutive *Fus ^ΔNLS/+^* mice. Germ line transmission of the recombinant allele was obtained and mice heterozygous and homozygous for the targeted allele will hereafter be referred to as *Fus^exon15^*. We first sought to validate the functionality of the new allele. To this end, primary cortical neurons of homozygous *Fus^exon15^* mice were transduced with CRE using an adenoviral vector (Ad-CRE). We observed Ad-CRE dose-dependent recombination of the *Fus* locus (**Figure S1B**), as well as decreased immunoreactivity to NLS FUS antibody (**Figure S1C**). To specifically mislocalize FUS in adult projection neurons, we crossed *Fus^exon15^* mice with transgenic mice expressing tamoxifen-inducible CRE specifically in excitatory projection neurons (Thy1^CRE-ERT2^ mice (Young et al., 2008)). These mice also express YFP from the same construct allowing the unambiguous identification of targeted cells (Young et al., 2008). Through two cross-breeding steps we generated Thy1^CRE-ERT2^/ *Fus^exon15^* mice, either homozygous or heterozygous for *Fus^exon15^.* Control mice were littermate *Fus^exon15^* homozygous mice, devoid of Thy1^CRE-ERT2^ transgene. All mice underwent 5 days of Tamoxifen gavage at 6 weeks of age and molecular analyses were performed 10 weeks after the induction of recombination (**Figure 3A**). Genotyping showed that a fraction of the *Fus* allele was efficiently recombined (**Figure S2A**). This recombination led to increased total *Fus* mRNA (**Figure S2B)** and mutant *Fus* mRNA as detected using RT-qPCR (**Figure S2C**). Moreover, nuclear/cytoplasmic fractionation followed by western blotting showed decreased immunoreactivity of FUS using an antibody targeting the C-terminal NLS, as well as increased cytoplasmic levels of FUS using an antibody recognizing the N-terminal domain in cortical extracts of Thy1^CRE-ERT2^/ *Fus^exon15^* mice (**Figure 3B-C**). Moreover, we observed increased cytoplasmic FUS staining when using an antibody targeting N-terminal FUS (**Figure 3D**) while FUS NLS immunoreactivity disappeared in YFP-positive (hence recombined) cells in the frontal cortex (**Figure S3A,** quantified in **Figure S3B**). Lastly, Thy1^CRE-ERT2^/ *Fus^exon15^* cortex showed hallmarks of FUS dysfunction as illustrated by an upregulation of *Taf15* (**Figure S4A**), as well as decreased inclusion of introns 6 and 7 of the *Fus* gene or of *Sfpq* intron 9 (**Figure S4B-D**), all of which are previously documented molecular alterations downstream of FUS mislocalization (Humphrey et al., 2020; Luisier et al., 2018; Sanjuan-Ruiz et al., 2021; Tyzack et al., 2019; Tyzack et al., 2021). As a whole, these results show that the recombination was efficient, and led to the mislocalization of FUS selectively in CPNs.

**Figure 3:**
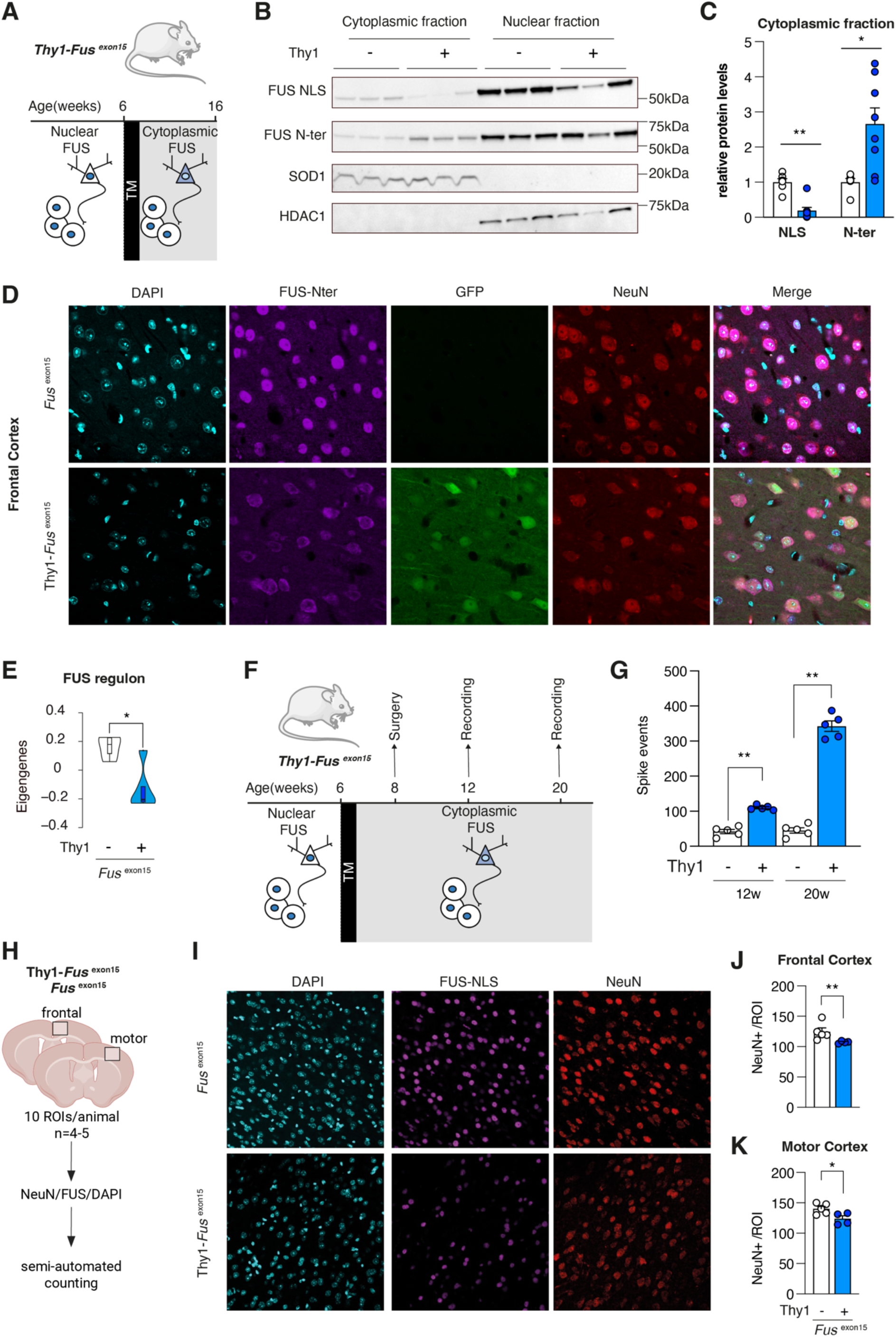
molecular, histological and electrophysiological consequences of adult mislocalization of FUS in cortical projection neurons. **A.** Scheme of the experimental design. Cortical projection neurons show normal nuclear FUS until tamoxifen administration (TM) at 6 weeks of age. After this age, FUS is permanently mislocalized in the cytoplasm of these neurons, but not in other cell types. **B.** Representative immunoblot analysis of FUS protein extracted from the frontal cortex, predicted molecular weights (MW) in kilodalton (kDa) are shown on the left. SOD1, mostly found in the cytoplasm, and HDAC1 exclusively found in the nucleus, were used as control of the purity of extracts. Two antibodies for FUS, targeting either the N-terminal part of FUS or the NLS (in C-terminal) were used. **C.** Quantification of FUS immunoreactive bands in cytoplasmic fractions relative to control mice (two-tailed Mann-Whitney test: *p=0.0127 and **p=0.0013). **D.** Representative widefield images of the immunofluorescent staining of cortical neurons in the frontal cortex of 4-month-old mice. GFP and NeuN staining was used to identify neurons (NeuN) recombined (GFP). The FUS antibody used here targets the N-terminal part of FUS, we observed a clear FUS cytoplasmic mislocalization in cortical neurons of Thy1^CRE-ERT2^/ Fus^exon15^ mice. **E.** FUS regulon eigengene expression in prefrontal cortex of Thy1^CRE-ERT2^/ Fus^exon15^ and control mice 3 months after TM (two-tailed t-test: T=-3.6, *p=0.013. **F.** Scheme of the experimental design for ECoG recording. **G.** Assessment of spikes events (defined as an event higher than a window discriminator of 4 times the baseline standard deviation calculated from control mice) at 12 and 20-weeks of age in Thy1^CRE-ERT2^/ Fus^exon15^ and control mice. Thy1^CRE-ERT2^/ Fus^exon15^ mice show a number of events two times greater than control mice at 12 weeks (two-tailed Mann-Whitney test; **p=0.0079), and five times greater at 20 weeks two-tailed Mann-Whitney test; **p=0.0079). Cortical activity alteration in transgenic mice significantly increases over time (two-way ANOVA: F_time_ =146.7, p<0.0001) depending on the genotype (two-way ANOVA: F_genotype_ =455.4, p<0.0001). **H-K.** Neurodegeneration characterization. **H.** 10 slices per mouse in the frontal cortex or in the motor cortex were selected for counting. **I.** Representative widefield images of the immunofluorescent staining of cortical neurons in the frontal cortex of 4-month-old mice. NeuN staining was used to identify and count neurons in the upper layers (mainly layer 2-3 and 5). **J.** We highlighted a significant decrease in the number of neurons in the frontal cortex of the transgenic mice (nested t-test: ** p=0.0086) and **K.** in the motor cortex of those (nested t-test: **p=0.0242). Results are expressed as mean ± SEM, control mice are represented in black and Thy1^CRE-ERT2^/ Fus^exon15^ in blue. **C.** Study of FUS mislocalization, n = 6 for control mice and n = 8 for Thy1^CRE-ERT2^/ Fus^exon15^. **E.** Study of FUS regulon. n = 5 for control mice and n = 5 Thy1^CRE-ERT2^/ *Fus^exon15^* mice (t-test: *p=0.013). **F.** Study of cortical activity, n = 5 for control mice and n = 5 Thy1^CRE-ERT2^/ *Fus^exon15^* mice. **I-K.** Study of neurodegeneration, n = 5 for control mice and n = 4 _Thy1CRE-ERT2/ *Fus*_*_exon15_* _mice._

Having established an animal model with isolated FUS mislocalization in adult CPNs, we first sought to determine whether this intervention recapitulated the gene expression fingerprint observed in both patients and *Fus*^ΔNLS/+^ mice. We performed high-depth bulk RNA sequencing in the frontal cortex of Thy1^CRE-ERT2^/ *Fus^exon15^* mice and control mice 2.5 months after recombination, which showed strong downregulation of the FUS regulon eigengenes (**Figure 3E**). Beyond the FUS regulon eigengenes, we identified 1127 differentially expressed (DE) genes between Thy1^CRE-ERT2^/ *Fus^exon15^* mice and controls (FDR <0.05) (**Figure S5A**) that were mostly related to neuronal projection and synapses (**Figure S5B-C**). Consistent with altered synaptic and electrical activity, Thy1^CRE-ERT2^/ *Fus^exon15^* mice developed increased spike event activity as recorded upon longitudinal electrocorticography at 12 and 20 weeks of age (i.e. 6 and 14 weeks after tamoxifen) (**Figure 3F-G**). We then counted NeuN-positive neurons 10 weeks after recombination in both frontal and motor cortex using semi-automated counting procedures (**Figure 3H**) and observed a significant loss of neurons in both regions (**Figure 3I-K**). Thus, adult-onset mislocalization of FUS triggers molecular and electrical alterations, as well as neuronal loss observed in ALS patients.

### Adult-onset FUS mislocalization in cortical projection neurons phenocopies ALS-ci and ALS-bi

We did not observe weight loss or decreased motor performance in the inverted grid and grip tests conducted in Thy1^CRE-ERT2^/ *Fus^exon15^* mice (**Figure S6A-D, Figure S7A**). Consistent with preserved motor function, lower motor neurons counts were unaltered despite efficient FUS mislocalization in these cells (**Figure S6E-G**). As ALS is also associated with impaired cognitive and behavioral function in ALS-ci and ALS-bi patients(Abrahams, 2023), we evaluated executive functions in Thy1^CRE-ERT2^/ *Fus^exon15^* mice (**Figure 4A**). We observed that Thy1^CRE-ERT2^/ *Fus^exon15^* mice displayed a lower mean score and a higher number of untorn cotton pieces when building their nests as compared to control mice (**Figure 4B-C**), suggesting that the truncation of FUS in adult CPNs of mice leads to difficulties in planning, or planning and executing the successive steps involved in nesting. To assess the general sociability and interest in social novelty of Thy1^CRE-ERT2^/ *Fus^exon15^* mice, we then performed the three-chamber test six weeks after induction of recombination. We first assessed the social preference of mice by presenting them a cage containing a mouse and an empty cage, and by measuring the time spent touching and sniffing them. We observed, as expected, that control mice spent significantly more time with the mouse (**Figure 4D**, solid black line) than with the empty cage during the first two trials (**Figure 4D**, dotted black line). This was also true for Thy1^CRE-ERT2^/ *Fus^exon15^* mice but they showed significantly less interest that the control mice (**Figure 4D**, solid *versus* dotted blue lines). We then assessed social memory of mice by presenting them a known mouse (i.e. the one already met during the social preference phase of the test) and a novel mouse. We observed that the contacting time of Thy1^CRE-ERT2^/ *Fus^exon15^* mice with the novel mouse was significantly lower compared to that of control mice (*p* < 0.001). As a consequence, the recognition index was significantly decreased (towards chance level) compared to that of control mice (**Figure 4E-F**). These results were replicated in another paradigm, the resident intruder test (**Figure 4G-H**), thus highlighting impairments in social functions, and in particular decreased social interest, in Thy1^CRE-ERT2^/ *Fus^exon15^* mice. Contrastingly, adult neuronal FUS mislocalization was not sufficient to trigger general memory defects as evidenced by an unaltered short-term or long-term object recognition (**Figure 4I-J**). Importantly, these mice did not suffer from generalized anxiety (**Figure S7B-D**) or lack of motivation (**Figure S7E-G**) that could explain sociability alteration. Thus, adult onset mislocalization of FUS in CPNs selectively impairs executive function, social interaction and social memory but preserves motor function or novel object recognition task. In all, adult onset mislocalization of FUS in CPNs provides a mouse phenocopy of ALS-ci/bi patients in the absence of motor symptoms.

**Figure 4:**
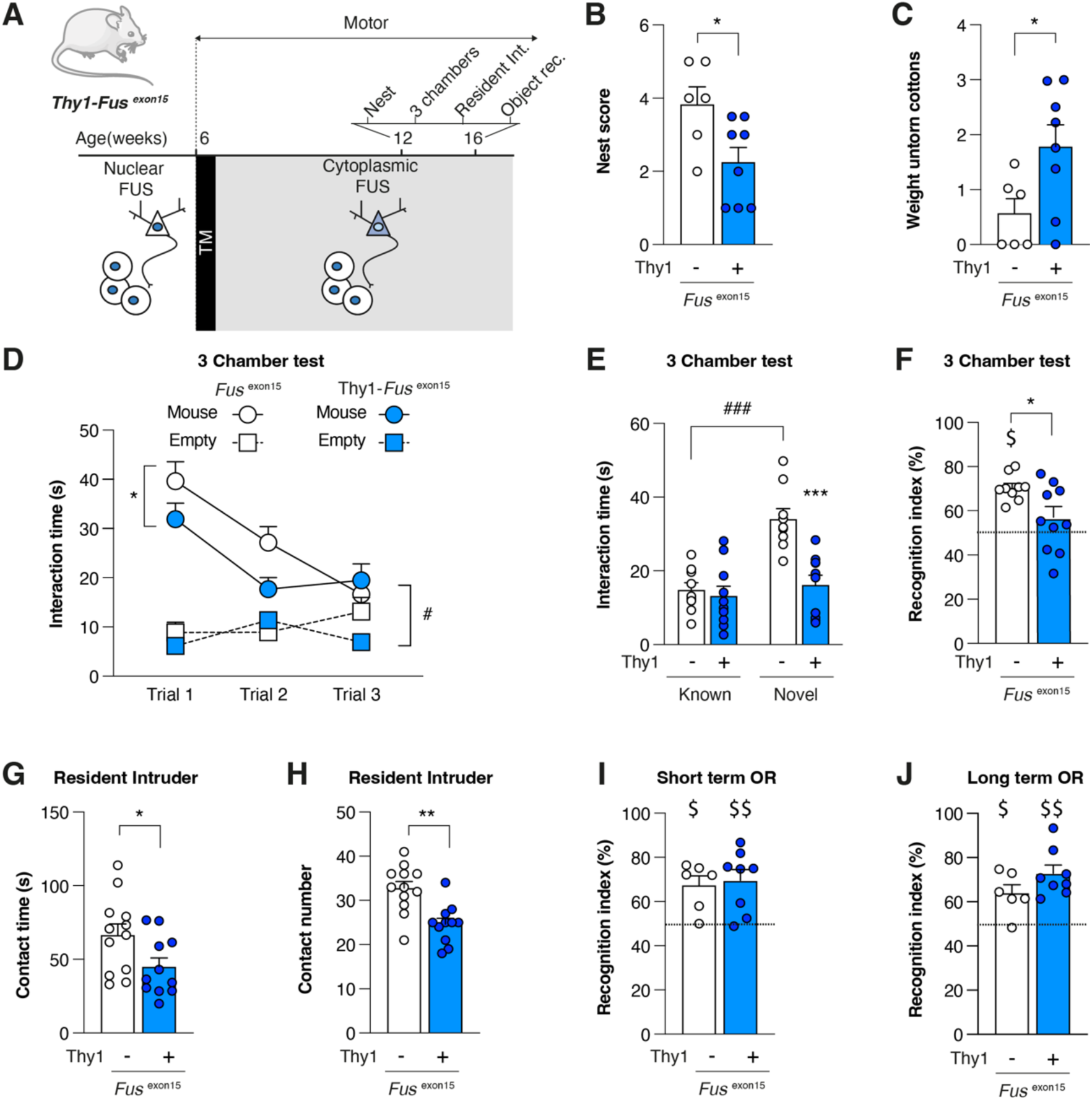
FUS mislocalization in adult neurons leads to ALS-related behavioral and cognitive impairment. **A.** Scheme of the experimental design. **B-C**. Nesting abilities assessment of Thy1^CRE-ERT2^/ Fus^exon15^ and control mice. **B.** Score attributed to the nests built in a 24hours period starting with 3 g of pressed cotton pieces. We observed that transgenic mice construct nest that are less complicated that the one performed by control mice (two-tailed Mann-Whitney test: *p=0.030). **C.** Mass of pressed cotton pieces which were found untorn after 24h. The mass of untorn cotton pieces was significantly higher for Thy1^CRE-ERT2^/ Fus^exon15^ than for control mice (two-tailed Mann-Whitney test: *p=0.045), highlighting difficulties to plan and/or execute the successive steps involved in nesting. **D-F** Sociability and social memory assessed using 3 chamber test. **D** During the first 3 trials, we measured the time spent contacting an empty cage or a cage with a mouse (mouse) while during the fourth trial, an unknown mouse is introduced (novel mouse) and allowed to assess the innate attractivity of mouse for novelty. Three-way ANOVA analysis showed a decrease of interest over time (F_time_ =8.217, p=0.0008), a higher interest for the mouse than the empty box (F_boxes_ =157.6, #p<0.0001) and a difference between genotype (F_genotype_ =4.075, *p=0.0488). **E.** Social memory evaluation by presenting a mouse already met (during trial 1-3, i.e. Known mouse) and a novel mouse. The contacting time of Thy1^CRE-ERT2^/ Fus^exon15^ mice with novel mice was significantly lower compared to those of control mice (two-way ANOVA: F_genotype_=14.39, ***p=0.0006). Šídák’s multiple comparisons test indicated significant higher exploration of novel mice in comparison to the known one in control mice (### p<0.0001). This is not the case for Thy1^CRE-ERT2^/ Fus^exon15^ mice (p=0.8783). **F.** The recognition index, computed as the contacting time with novel mouse divided by the total contacting time, was significantly decreased compared to those of control mice (two-tailed Mann-Whitney test: *p=0.035). When we compared this recognition index to chance level (placed at 50 and indicated by the horizontal dotted line), we observed that only control mice presented an index higher than chance level (one sample t-test: control mice $p<0.0001 and Thy1^CRE-ERT2^/ Fus^exon15^, p=0.2233). **G-H.** Social alteration assessed in another paradigm, resident intruder test. We placed in the tested mouse cage an intruder and assess for 5 min the total contacting time (**G**) and the total number of contact (**H**). We confirmed the social alteration observed in Thy1^CRE-ERT2^/ Fus^exon15^ as they spent less time contacting the intruder than the control mice (two-tailed Mann-Whitney test: *p=0.0317) and showed a lower number of total interactions (two-tailed Mann-Whitney test: **p=0.0011). **I-J.** Short-term (10 min) and long term (24hours) object recognition memory. **I.** We did not observe any short-term object memory alteration (two-tailed Mann-Whitney test: p=0.662). Both groups spent more time than chance level (indicated by the horizontal dotted line) exploring the novel object (one sample t-test: $p=0.011 for control mice and $$p=0.0058 for Thy1^CRE-ERT2^/ Fus^exon15^ mice). **J.** Long-term (10 min) object recognition memory is not affected either (two-tailed Mann-Whitney test: p=0.345). Both groups spend more time than chance level exploring the new object ($p=0.016 for control mice and $$$p=0.0005 for Thy1^CRE-ERT2^/ Fus^exon15^ mice). Results are expressed as mean ± SEM, control mice are represented in black and Thy1^CRE-ERT2^/ Fus^exon15^ in blue. **D-F**. Study of 3 sociability in 3 chamber test, n = 9 for control mice and n= 10 for Thy1^CRE-ERT2^/ Fus^exon15^ mice. **G-H** Study of sociability in resident intruder test, n= 12 control mice and n = 11 for Thy1^CRE-ERT2^/ Fus^exon15^ mice. **I-J** Study of object recognition, n = 6 control mice and n = 8 for Thy1^CRE-ERT2^/ Fus^exon15^ mice.

### FUS pathology and downregulation of the FUS regulon in patients with ALS-ci

Our results in mice provided a causal link between cytoplasmic FUS mislocalization and cognitive and behavioral impairment. We then sought to determine whether this relation could be observed in ALS-ci patients. To this aim, we selected 6 deeply clinically phenotyped *C9ORF72* cases with post-mortem tissue paired with regional cognitive scoring performed for that functional domain. We selected BA39 as a language processing brain region that had been assessed during life using the Edinburgh Cognitive ALS Screening (ECAS) tool. Cases were stratified into ALS-ci (ECAS language domain score < or = 26) and ALS (ECAS language domain score >26) based on published diagnostic cutoffs. Cases with ALS-ci exhibited evidence of FUS pathology (cytoplasmic and nuclear pathology) (**Figure 5A**), which was less frequently encountered in ALS cases that were cognitively intact (**Figure 5B-C**). Indeed, FUS pathology, quantified using digital burden scoring (Rifai et al., 2022) correlated with ECAS score in this region scale (**Figure S8**) demonstrating that the burden of FUS pathology, measured in a non-biased way, correlates with cognitive function in these cases. To provide orthogonal evidence of altered FUS function in ALSci, we re-analyzed our dataset of snRNAseq in ALS and ALS-ci patients in frontal regions BA44 and BA46 (Petrescu et al., 2025) and merged this dataset with previously published BA4 and BA9 regions involved in motor control (Pineda et al., 2024). We identified 287,275 excitatory neurons and, among those 1,547 L5-ET neurons, that expressed typical L5-ET expression markers such as *ADRA1A, POU3F1,VAT1L and SULF2* (**Figure 5D-E**). Merging the two datasets revealed two L5-ET subtypes, with their own expression markers (**Figure 5E**) and different distributions across cortical regions studied (**Figure 5F**). Importantly, we observed a diminished proportion of L5-ET neurons, but not L5-IT neurons, in ALS, and even more in ALSci patient (**Figure 5G-H**) in these datasets, further confirming the selective involvement of this neuronal population in ALS and ALSci. Importantly, expression of the FUS regulon was significantly downregulated in L5-ET and L5-IT neurons of ALSci patients as compared to controls (**Figure 5I**). ALS patients showed an intermediate situation with decreased expression of the FUS regulon only in L5-IT neurons (**Figure 5I**). This relationship between cognitive impairment and FUS regulon expression in L5-ET neurons was also observed at the individual patient level (**Figure 5J**) and there was a positive correlation between FUS regulon expression in L5-ET neurons and total ECAS scores of studied patients (**Figure 5K**). Strikingly, this relationship was mostly observed in BA46, involved in executive function, as compared to BA44 (**Figure 5L**). Last, we sought to determine whether cognitive function and FUS dysregulation could be related to an accumulated genetic burden of dementia. To answer this question, we processed whole genome-sequencing of patients with ALS and ALSci (Petrescu et al., 2025), and filtered out gene variants involved in dementia to compute a genetic burden that could be correlated with FUS regulon expression and cognitive function (**Figure 5M**). Indeed, a high genetic burden (>2 missense variants) was associated with decreased ECAS score (**Figure 5N**) and, at the individual patient level, ALSci patients showing a higher genetic burden (**Figure 5J**). Accumulation of deleterious missense variants led to decreased cognitive performance in executive, language or memory subscores (**Figure 5O**). Consistently, expression of the FUS regulon was negatively correlated to genetic burden of dementia (**Figure 5P**). In all, our results suggest that ALSci develops in ALS patients with increased genetic burden of dementia, leading to cytoplasmic FUS mislocalisation and decreased expression of FUS downstream target genes in L5-ET vulnerable neurons.

**Figure 5:**
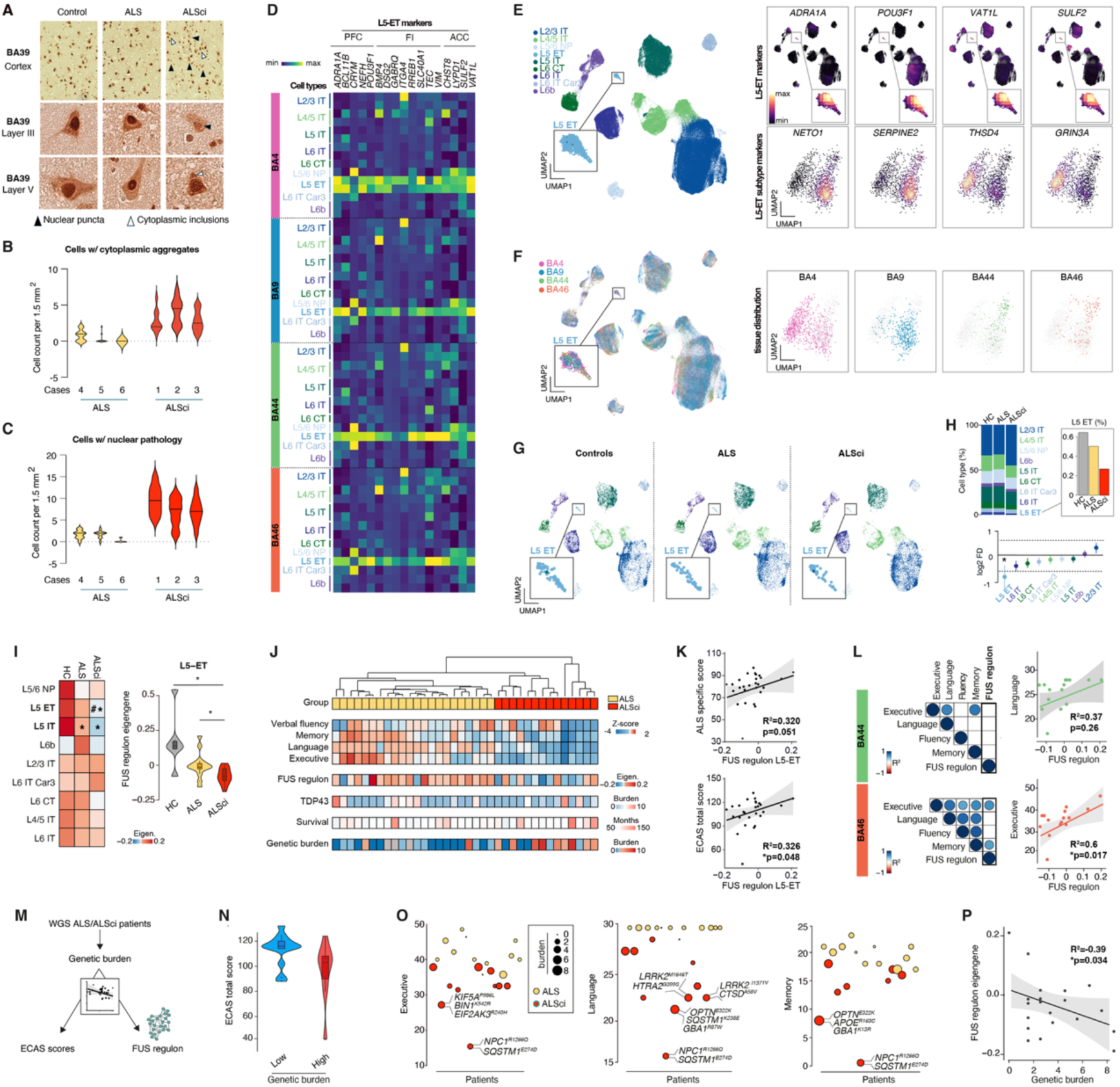
FUS mislocalization and downregulation of FUS regulon in vulnerable neurons of patients with ALS and cognitive impairment. **A.** Representative photomicrographs of *C9ORF72* human post-mortem tissue taken at 10x (top panel) and 40x magnification with optical zoom (middle and lower panels) demonstrating DAB immunohistochemical staining for FUS protein in BA39 (language brain region). These cases had undergone ECAS testing during life, evaluating language function of this brain region, and based on clinically approved cut offs cases were classified as affected (ECAS score < or = 26) or unaffected (ECAS score > 26). Black arrowheads indicate nuclear pathology, and white arrowheads indicate cytoplasmic pathology within layers III and V of the cortex. **B-C.** Cell counts were performed by counting the number of cells affected by cytoplasmic pathology (B) or nuclear pathology (C) in 10 randomly assigned regions of interest (1.5 mm2) in three cases for each group. Cell counts were performed by a pathologist who was blinded to case and demographic data. ALS cases with normal language function are plotted in yellow and with language dysfunction plotted in red. Each case is plotted demonstrating the variability between and within cases. **D.** Heatmap showing cell-type specific expression of L5-ET marker genes in 9 excitatory neuron subclasses spanning four different tissues (BA4,BA9,BA44 and BA46). **E.** Uniform manifold approximation and projection (UMAP) UMAP plot projecting gene expression from 287,275 excitatory neurons colored by subclasses and density plot showing the expression profile of L5-ET top marker genes (*ADRA1A,POU3F1,VAT1L* and *SULF2*) in L5-ET subgroups and L5-ET subtypes top marker genes (*NETO1, SERPINE2, THSD4* and *GRIN3A*) **F.** UMAP plot colored by sampled tissue show selective regional presence of L5-ET subgroups across motor areas (BA4) and prefrontal area of the cortex (BA9, BA44, BA46). **E.** UMAP plot split according to diagnosis show a reduction in L5-ET neurons. **H.** Bar plot showing proportion of each excitatory neurons subclass sampled across all donors and showing a reduction in L5-ET neurons in ALSci patients compared to ALS and HC (inset on the right). The lower panel shows results of a permutation-based test identifying L5-ET neurons as significantly depleted in ALSci patients (FDR<0.05 & log2 Fold difference <-0.58). **I.** Heatmap displays FUS regulon expression across major subclasses of excitatory neurons in controls (HC), ALS and ALSci patients. A significant decrease is observed in L5-ET neurons in ALSci compared to controls (Wilcoxon rank sum test Bonferroni adjusted **p=0.007) and in ALS compared to ALSci patients (Wilcoxon rank sum test Bonferroni adjusted **p=0.044). *Adjusted-p <0.05 vs HC, # Adjusted-p <0.05 vs ALS. The violin plot on the right shows FUS regulon expression difference in L5-ET neurons of ALSci compared to controls (Wilcoxon rank sum test Bonferroni adjusted **p=0.002) and ALS (Wilcoxon rank sum test Bonferroni adjusted *p=0.029) but not between ALS and HC (Wilcoxon rank sum test Bonferroni adjusted p=0.051). **J.** Unsupervised clustering of ALS donors (n=36) screened for cognitive impairment shows a clear separation between ALS patients (n=22) and ALSci patients (n=14) and heterogeneity in clinical scores. Heatmap displays clinical score across 4 ECAS domains that are ALS specific (Executive,Language and Verbal Fluency) and non-specific (Memory). Overlay heatmaps show FUS regulon expression across donors, TDP43 burden, disease duration and genetic burden of rare and common variants associated with neurodegenerative disease-associated dementia. **K.** Scatter plot shows FUS regulon expression correlation with clinical ALS specific score (R2=0.32,p=0.051) and ECAS total score (R2=0.326,*p=0.048). **L.** Corrplot displays strong correlation across clinical subscore across individuals and a significant correlation of the FUS regulon expression with executive function in BA46 (R2=0.60,*p=0.017) but not with language in BA44 (R2=0.37,p=0.26) illustrated in scatter plots on the right. **M.** Strategy to relate genetic burden of dementia to FUS regulon expression and cognitive impairment. **N.** Violin plot shows a significant reduction of the ECAS total score in ALS patient with high genetic burden (>=2 screened missense variants per patient) vs low (< 2 screened missense variants per patient) (Wilcoxon rank sum test *p=0.034). **O.** Dot plots across patients colored by diagnosis shows an increase burden of the number of genetic variation observed per individual in known genes and its association with clinical subscores. **P.** Scatter plot shows FUS regulon expression inverse correlation with genetic burden across donor (R2=-0.39,*p=0.034).

### The FUS regulon is enriched in ALS genetic risk and allows for prioritizing novel ALS-ci/bi genes

Our previous results provided indirect evidence that FUS and its regulated genes might lie in the same genetic pathway than other dementia related genes. Interestingly, the heritability of ALS is enriched in FUS binding sites (Megat et al., 2023). We thus hypothesized that the FUS regulon could be enriched in ALS-risk genes, and we applied stratified linkage disequilibrium (Bulik-Sullivan et al., 2015) (LD) score regression (S-LDSC) to the most recent ALS genome-wide association study (GWAS)(van Rheenen et al., 2021) (**Figure 6A**). We compared ALS heritability estimates in FUS regulon gene to a randomly sampled gene set over 10 iterations and used ALS GWAS data as a positive control. As shown in **Figure 6B**, FUS regulon genes are enriched in ALS heritability (E=87.7 fold, *Bonferroni p < 0.05). As expected, we observed a strong enrichment of the ALS GWAS (E=387.3 fold, *Bonferroni p < 0.05) but no enrichment in the randomly sampled gene set (Bonferroni p > 0.05). These results suggest that the FUS regulon is highly enriched in ALS heritability and might contain new potential ALS genes. Then, we sought to refine our analysis through the comparison of the FUS regulon with genes known to cause ALS, ALS-FTD or pure FTD. When overlapping genes in the FUS regulon with a curated list of ALS genes, we observed a significant enrichment of genes involved in ALS-FTD, including (*FUS*, *KIF5A, C9ORF72*, *ERBB4*, *HTT* or *GBA*) (Fisher’s exact test; p=2.2.10^−06^) (**Figure 6C**).

**Figure 6:**
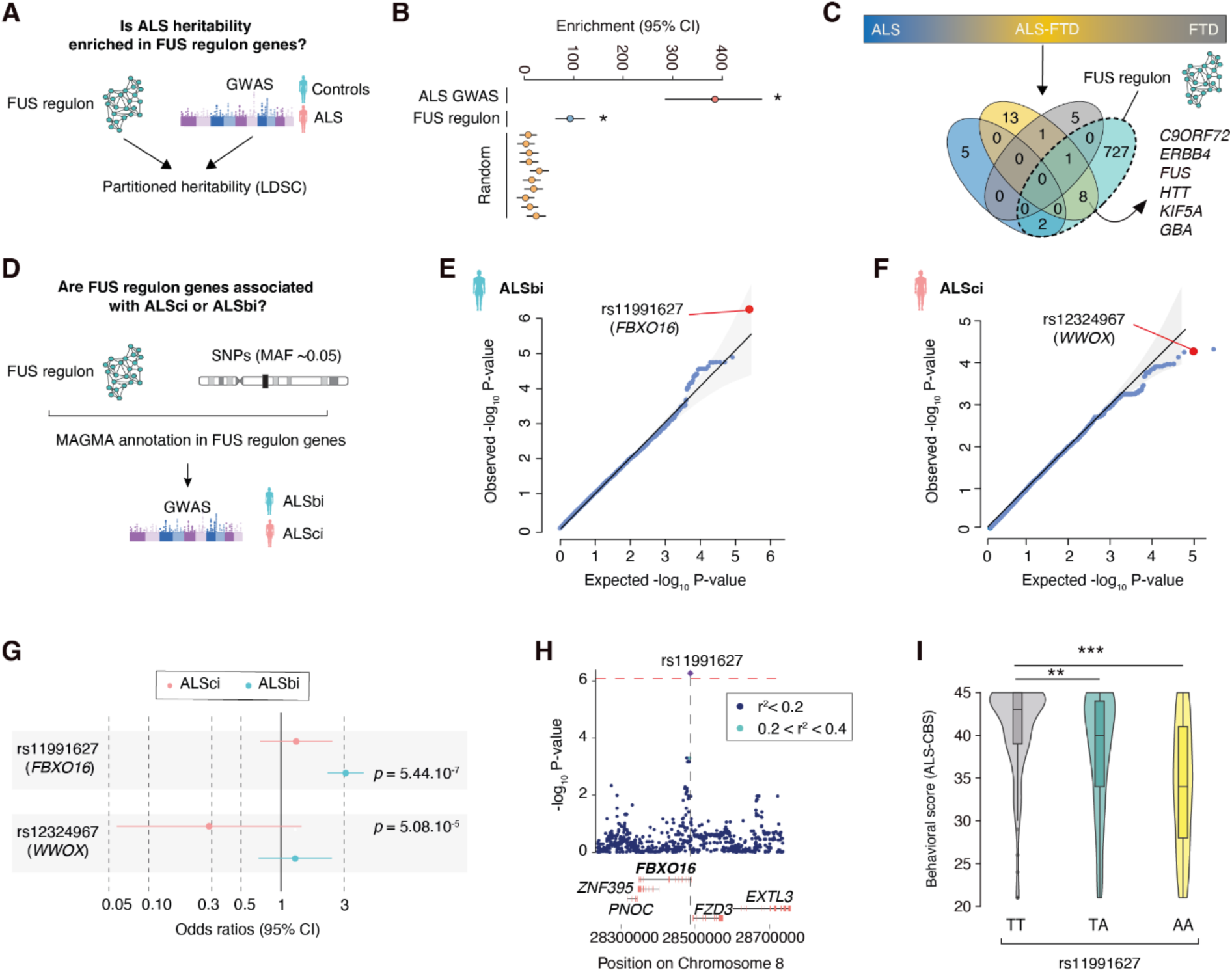
FUS regulon is enriched in ALS/FTD heritability and prioritizes FBXO16 as a novel risk factor for ALS associated with behavioral impairment. **A.** Strategy to characterize heritability through integration of GWAS with the FUS regulon. Each gene in the FUS regulon was mapped to the associated TSS and/or enhancer through single nuclei ATACseq (snATACseq) and linkage disequilibrium score regression computed as described previously **B.** Dots indicate enrichment and 95% confidence interval computed from LD-score regression. Two-sided p-values were derived from linear regression in LDSC analysis. * Indicates significance after Bonferroni correction p<0.05/12. Results indicate enrichment for ALS-GWAS *p=1.07.10^−24^ and the FUS regulon *p=1.3.10^−9^ in ALS heritability but no significant enrichment for randomly samples set of genes (p>0.05). **C.** Genetic risk factors for ALS, ALS-FTD and FTD were overlapped with the FUS regulon. Venn diagram show overlap between each group with a significant overlap between ALS-FTD gene and FUS regulon (Fisher’s exact test p=2.2.10^−06^) but not with ALS only (Fisher’s exact test p=0.054) or FTD only (Fisher’s exact test p=0.33). **D.** Strategy to perform GWAS of ALS associated with cognitive (ALS-ci) or behavioral impairment (ALS-bi). Whole genomic sequencing from ALS patients were filtered for SNPs with minor allele frequency above 5% (MAF > 0.05). WGS were then filtered based on gene overlap with FUS regulon genes leading to 131,202 SNPs used for GWAS. **E.** Quantile–Quantile plot depicting on x-axis the −log10 of expected p value versus the actually measured p values from generalized logistic regression for 131,202 SNPs association with ALS-ci. Genome-wide correction for multiple testing was set at P < 1 × 10^−6^. Highlighted SNP in red reaches genome-wide significant (p=5.44.10^−07^) and is located in the FBXO16 gene. **F.** Quantile–Quantile plot depicting on x-axis the −log 10 of expected p value versus the actually measured p values from generalized logistic regression for 131,202 SNPs association with ALS-bi. Genome-wide correction for multiple testing was set at P < 1 × 10^−6^. Highlighted SNP in red reaches suggestive significance (p=5.08.10^−05^) and located in WWOX gene. **G.** Forest plot display per-GWAS association of rs11991627 (*FBXO16*) and rs12324967 (*WWOX*) in ALS-ci and ALS-bi. Odds ratio (OR) values and 95% CI for each cohort are depicted in different color. The graphs display the means and 95% confidence interval. **H.** Locus zoom plot showing the SNP (+/− 500KB) rs11991627 (*FBXO16*) association with ALS-bi. Red dashed lines show the genome-wide significant SNP at a p-value < 6.10^−07^ and colored dots represent LD with the lead variant (red diamond). **I.** Violin plots shows behavioral score in rs11991627 carriers of homozygous reference allele (n=139) compared to heterozygous carriers (n=104) (One-way ANOVA F_genotype_=14.26, p=1.31.10^−06^, Tukey’s post-hoc **p=2.58.10^−03^) and to homozygous carriers (n=25) (Tukey’s post-hoc ****p=5.67.10^−06^).

Having shown that FUS regulon expression is altered in ALS-ci and is enriched in ALS-FTD risk genes we sought to test whether the FUS regulon could help us prioritizing new genetic risk factors associated with either ALS-ci or ALS-bi. To do so, we analyzed whole genome sequences (WGS) from 546 ALS patients of the Answer ALS cohort who have been evaluated for cognitive and behavioral impairment through questionnaire screening according to ALS cognitive Behavioral Screen (ALS-CBS) scale (**Figure 6D**) (Baxi et al., 2022).WGS SNPs were filtered to map FUS regulon genes resulting in a final set of 131,202 SNPs falling onto at least one gene of the FUS regulon. Interestingly, we observed that rs11991627 located in the *FBXO16* gene is significantly associated with the risk of ALS-bi (Logistic regression: *OR = 3.22 (1.94-4.23) p*=5.44.10^−07^) while it does not associate with ALS-ci (Logistic regression: *OR = 1.30 (0.16-2.45) p*=0.098) (**Figure 6E-H**). Conversely, we observed a suggestive association of rs12324967 in the *WWOX* gene, previously associated with cognitive impairment in Parkinson’s disease (Liu et al., 2021), with ALS-ci (Logistic regression: *OR = 0.28 (0.03-1.43) p*=5.08.10^−05^) (**Figure 6E-H**). The rs11991627 allele in *FBXO16* shows an additive effect on behavioral score (One-way ANOVA: \*\*\**p=1.31.10^−06^*), suggesting that *FBXO16* rs11991627 carriers have increased risk of behavioral impairments associated with diagnosed ALS (**Figure 6I**). These results show that the FUS regulon includes novel genes associated to cognitive and/or behavioral impairment in ALS, including *FBXO16* or *WWOX*.

### *FBXO16* PTV carriers show behavioral abnormalities, fronto-temporal atrophy and increase expression of fluid biomarkers associated with dementia

We then sought to independently validate whether variants in *FBXO16* could represent risk factors for ALS-FTD related cognitive impairment. To this end, we leveraged large-scale UK Biobank data (Sudlow et al., 2015). In UK Biobank, we identified carriers of *FBXO16* protein-truncating variants (PTV) and tested whether these individuals displayed behavioral, imaging or protein biomarker changes associated with ALS and/or FTD (**Figure 7A**). We identified 231 *FBXO16* PTV carriers (n=231), and observed increased risk associated with behavioral and emotional states in these individuals (R45 category: *Bonferroni corrected-p < 0.05) (**Figure 7B**). The R45 subcategory of symptoms reported in the UK-Biobank dataset includes apathy, social isolation and irritability, and we observed that *FBXO16* PTV carriers showed increased risk of hostility behaviors (logistic regression: *Bonferroni corrected-p < 0.05), irritability (logistic regression: *Bonferroni corrected-p < 0.05) and nervousness (logistic regression: *Bonferroni corrected-p < 0.05). Moreover, we also observed a suggestive association with apathic behavior (logistic regression: p=0.018) (**Figure 7B**). These results suggest that carrying a *FBXO16* PTV variant leads to behavioral abnormalities generally associated with ALS-bi.

**Figure 7:**
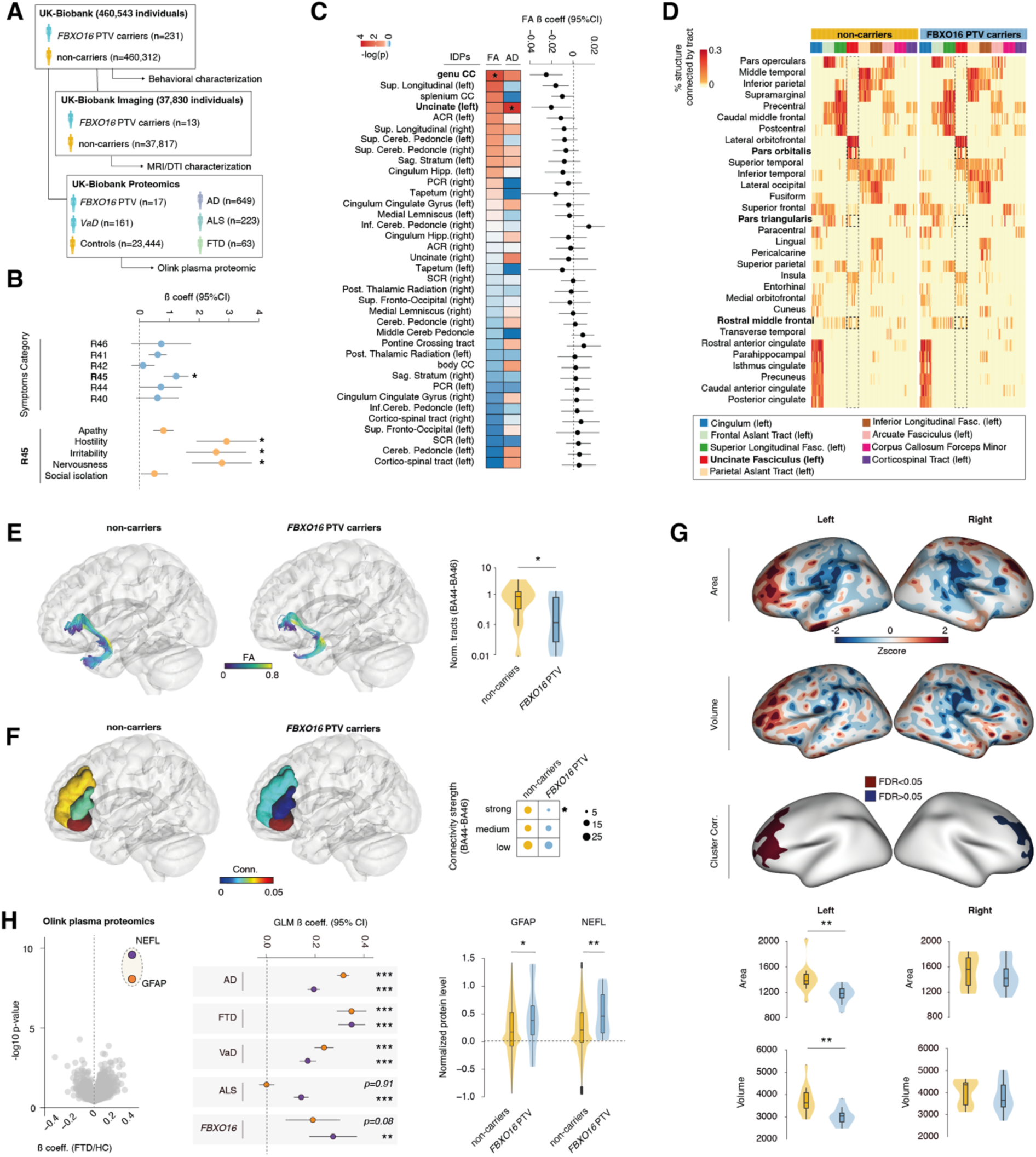
FBXO16 PTV carriers have behavioral deficits associated with fronto-temporal atrophy and increase plasmatic expression of NEFL and GFAP. **A** Strategy to characterize *FBXO16* PTV carriers in the UK Biobank, and numbers of individuals included. *FBXO16* PTV carriers (n=231) were screened for symptoms associated with cognitive or behavioral deficits among 460,312 individuals of European ancestry. We then evaluated alteration of the brain structure using diffusion and T1 structural MRI from FBXO16 PTV carriers (n=13) and 37,817 non-carriers controls individuals. Finally, we have used large-scale proteomics data from the UK-Biobank to test whether *FBXO16* PTV carriers (n=14) show significant alteration in major biomarkers when compared to age and sex-matched controls (n=21,604). Levels of different proteins were compared to incident diseased cases diagnosed with Alzheimer’s dementia (n=642), ALS (n=198), FTD (n=70) and Vascular dementia (n=161). **B.** Logistic regression analysis testing whether *FBXO16* PTV carriers’ status predict UK-Biobank associated symptoms and signs involving cognition, perception, emotional states and behavior. Forest plot shows significant association of *FBXO16* genotype with R45 category (Logistic regression Beta coefficient = 1.22; (0.81-1.64), *p=0.0030). *Significance is set at p-values < 0.05 / 6. A significant association is observed between *FBXO16* genotype and two R45 subcategory which are hostility (Beta coefficient = 2.92, (1.91-3.92), *p=0.0037) and nervousness (Beta coefficient = 2.76, (1.75-3.77), *p=0.006) and a suggestive association with apathic behavior (Beta coefficient = 0.80, (0.46-1.14), p=0.018). *Significance is set at Bonferroni corrected p-value < 0.05 / 6 = 0.008. **C.** Heatmap depicts −log10(p) obtained from generalized linear model testing whether FBXO16 genotype is a predictor of IDPs obtained from diffusion MRI imaging of FBXO16 carriers (n=13) and non-carriers (n=37,817). GLM p-values are depicted for the fractional anisotropy and axial diffusivity for both hemispheres and each of the white matter tracts. A significant association is observed between FBXO16 genotype and a decrease FA of the genu of the corpus callosum (Beta = −0.03 (−0.048,-0.012),* adjusted-p =0.041) and a decrease AD of the left uncinate fasciculus (Beta coefficient = −6.17.10^−05^ (−9.53.10^−05^, −2.8.10^−05^), *adjusted-p=0.337). **D-F.** Heatmap shows the percentage of each of the 31 structures from the Desikan-Killiany cortical parcellation atlas which is connected to the nine major white matter tracts in FBXO16 PTV carriers and non-carriers. We observe a decrease in the percentage of the structure connected through the left uncinate fasciculus to the pars orbitalis (BA47), the pars-triangularis (BA45) and the rostral-middle frontal (BA46). Dashed rectangles. The number of tracts from the left uncinate fasciculus connecting to BA47, BA45 and BA46 is significantly decreased in FBXO16 carriers compared to non-carriers (unpaired t-test: t=2.16, *p=0.045). Also, we observed a lower proportion of highly connected tracts of the uncinate fasciculus to BA44, BA45, and BA47 in *FBXO16* PTV carriers when compared to non-carriers (Fischer’s exact test: X-squared = 4.576, df = 1, *p = 0.032). **G.** Illustration shows the Z-score differential gray matter area and volume. Unbiased brain-wide cluster correlation analysis reveals a significant decrease in left gray matter area located on the frontal region in the pars triangularis (BA45) and the rostral medial frontal region (BA46). Region-of-interest reveals a significant decrease in the left pars triangularis gray matter area (unpaired-t test: t=3.24, **p=0.0047) and volume (unpaired-t test: t=3.33, **p=0.0036). **H.** Volcano plot shows a significant association between increase GFAP (Beta coefficient = 0.332, adjusted-p=3.25.10^−05^) and NEFL (Beta coefficient = 0.337, adjusted-p=1.96.10^−06^) protein levels in the plasma with incidence of FTD. Forest plot display association between GFAP and NEFL levels with dementia subtypes which significant for Alzheimer’s disease (GFAP-Beta coefficient = 0.29, ***p=3.38.10^−49^; NEFL-Beta coefficient = 0.17, ***p=1.08.10^−23^), vascular dementia (GFAP-Beta coefficient = 0.22, ***p=5.45.10^−06^; NEFL-Beta coefficient = 0.16, ***p=8.32.10^−04^). Only a significant association of NEFL with ALS (NEFL-Beta coefficient = 0.14, **p=3.17.10^−03^) but not with GFAP (NEFL-Beta coefficient = −0.003, p=0.92) In FBXO16 carriers a significant association is observed for NEFL (NEFL-Beta coefficient = 0.27, **p=0.0047) and trend for GFAP (GFAP-Beta coefficient = 0.19, p=0.085). Violin plot show a significant increase in mean GFAP protein expression (D-cohen = 0.47; one-tailed *permuted-p = 0.027) and NEFL (D-cohen = 0.67; one-tailed **permuted-p = 0.0023) in FBXO16 PTV carriers.

Next, we sought to test whether *FBXO16* PTV carriers could show altered brain connectivity and structure, and used the diffusion MRI and T1 structural MRI datasets (Littlejohns et al., 2020; Miller et al., 2016). Using image-derived phenotypes (IDPs) from diffusion MRI, we performed an unbiased association analysis to test whether *FBXO16* PTV carriers show altered white matter structure evidenced by a decrease in fractional anisotropy (FA) or axial diffusivity. Indeed, *FBXO16* PTV carriers (n=13) showed a large decrease in FA in the genu of corpus callosum (GLM regression: FDR<0.05) and decreased axial diffusivity in the left uncinate fasciculus (GLM regression: FDR<0.05) when compared to non-carriers individuals (n=37,817) (**Figure 7C**). Interestingly, previous studies showed a loss of white matter in the genu of the corpus callosum in patients diagnosed with behavioral variant FTD(Elahi et al., 2017; Mahoney et al., 2015) and decreased FA in the left uncinate fasciculus in patient diagnosed with behavioral variant FTD (Mahoney et al., 2015) that correlate with symptoms severity (D’Anna et al., 2016). Using a whole-brain connectomics analysis and focusing on the left uncinate fasciculus, we observed that *FBXO16* PTV carriers display a lower number of white matter tracts (unpaired t-test;*p<0.05) connected to distinct frontal region such as the pars orbitalis (BA47), the pars triangularis (BA45) and the rostral medial frontal cortex (BA46) (*Fisher’s exact test;*p<0.05) (**Figure 7D-F**), that are all parts of the inferior frontal gyrus (Broca’s area) with crucial role in semantic tasks and working memory (Munoz-Neira et al., 2019). As *FBXO16* PTV carriers showed strong alteration in white matter integrity in a specific region of the left frontal cortex, we studied possible grey matter changes and analyzed T1-MRI scans from *FBXO16* PTV carriers and non-carriers. We observed that *FBXO16* PTV carriers showed a strong decrease in area and volume of the left frontal regions that correspond to the pars triangularis (BA45) (Cluster-correlation; FDR<0.05) (**Figure 7D-F**). Region-specific ROI analysis indicated a significant decrease of the area (unpaired-t-test; **p<0.01) and volume (unpaired-t-test; *p<0.05) in the left pars triangularis (BA45) region in *FBXO16* PTV carriers while no differences were observed in the right hemisphere (**Figure 7G**). Finally, we studied whether *FBXO16* PTV carriers show altered plasma protein biomarkers associated with neurodegenerative diseases. Using the prospective UK-Biobank Olink-based proteomic cohort (Sun et al., 2023), we selected individuals with incident neurodegenerative disorders including ALS and dementia. After quality control and inclusion of age and sex-matched individuals across diseases, we included 24,557 adults without ALS or dementia in the UK Biobank, along with 1,073 incident cases and a follow-up time of 8.6 years. During this follow-up, 884 (3.7%) incident dementia cases were identified, of which 118 occurred within 5 years, 350 within 10 years and 415 beyond 10 years. In this population, we observed increased plasma neurofilament light chain (NEFL) (*GLM p =1.96.10^−06^*) and glial fibrillary acidic protein (GFAP) proteins (*GLM p =1.96.10^−06^*) in FTD (**Figure 7H**). These two proteins were also altered in different dementia subtypes, including Alzheimer’s disease (Bonferroni corrected p <0.05) and vascular dementia (Bonferroni corrected p <0.05) (**Figure 7H**). In this population, there were also 223 patients diagnosed with ALS, of which 108 occurred within 5 years, 93 had incidents within 10 years and 22 had incidents over 10 years. Interestingly, plasma GFAP expression was unchanged in pure ALS (GLM p > 0.05) while NEFL was significantly increased (Bonferroni corrected p <0.05) in incident dementia, in line with previous studies (Guo et al., 2024; Katisko et al., 2021). Interestingly, *FBXO16* PTV carriers showed significantly elevated levels of GFAP protein (permuted-p<0.05) as well as NEFL (permuted-p<0.01) (**Figure 7H**). Altogether, our results show that loss of *FBXO16* is associated with behavioral, neuroimaging and plasma biomarker evidence of altered fronto-temporal network, consistent with prodromal ALS/FTD.

## Discussion

Neurodegenerative diseases are defined by three major characteristics: a core set of symptoms (progressive paralysis for ALS), loss of specific neuronal populations and a typical proteinopathy (TDP-43 for ALS). However, the clinical picture associated with a specific neurodegenerative disease can include additional symptoms in some patients. In ALS, for instance, a substantial fraction of patients develops cognitive impairments. In these ALS-ci patients, the relation between cognitive impairment and TDP-43 pathology is unclear (Bampton et al., 2024; Rifai et al., 2024; Suzuki et al., 2024). In most neurodegenerative diseases, proteinopathies are frequently mixed, and the occurrence of multiple pathologies is generally related to more severe cognitive impairment. However, the contribution of individual pathologies to the heterogeneity of symptoms is unclear (Forrest and Kovacs, 2024; Maldonado-Diaz et al., 2024). Furthermore, in ALS and FTD, FUS proteinopathy is generally overlooked as it may involve cytoplasmic mislocalization rather than aggregation (Rifai et al., 2022; Tyzack et al., 2019).

A first important result of this study is that FUS mislocalization in CPNs drives ALS-ci/bi-like impairments in ALS. This result is substantiated by correlative evidence in patients as FUS mislocalization in CPNs is observed in ALS-ci patients, but not in those without cognitive impairments. Consistently, the typical signature of FUS-related dysfunction was observed in L5-ET neurons of ALS-ci patients and related to cognitive impairment at the individual patient level. Whilst the absence or presence of TDP-43 pathology is associated with the absence or presence of cognitive dysfunction (Gregory et al., 2020), no protein misfolding pathology has previously been shown to correlate with the burden of symptoms, highlighting the importance of this finding with respect to FUS. Beyond correlative evidence, we provide functional, *in* vivo, evidence that mislocalization of FUS in adult CPNs is sufficient to drive cognitive and behavioral impairment in a novel mouse model. These behavioral abnormalities were observed a few weeks after the induction of FUS mislocalization and phenocopied cognitive and behavioral impairments in ALS-ci/bi patients. Contrastingly, these mice were spared of motor defects even after more than one year of follow up. This finding indicates that the occurrence of FUS mislocalization in CPNs might drive cognitive impairment in ALS patients already presenting the motor symptoms. Our current results suggest that FUS mislocalization should be systematically included in the pathological characterization of ALS-ci/bi patients, and, beyond in neurodegenerative diseases with cognitive and behavioral impairments.

How could FUS mislocalization be triggered in CPNs of ALS-ci/bi patients? In rare cases, ALS patients carry germline mutations of *FUS* that increase FUS mislocalization(Kwiatkowski et al., 2009; Vance et al., 2009). These mutations generally lead to severe motor phenotypes and there are few studies characterizing possible cognitive impairments in these *FUS*-ALS patients. However, learning disabilities and mental retardation have been observed in *FUS*-ALS patients especially in severely affected patients (Goldstein et al., 2022; Grassano et al., 2022; Hirayanagi et al., 2016; Lanteri et al., 2021). There are also reports of patients with *FUS* variants showing fronto-temporal dementia phenotype either isolated or with an initial presentation of ALS-FTD(Aguzzoli et al., 2022; Broustal et al., 2010; Van Langenhove et al., 2010). However, most ALS-ci patients do not present a germline *FUS* mutation, and FUS mislocalization is likely, in these cases, secondary to other pathogenic events. In cell models, FUS mislocalization can be elicited by various stresses including arsenite (Bosco et al., 2010; Dormann et al., 2010; Szewczyk et al., 2023), osmotic stress (Gao et al., 2022; Hock et al., 2018), antiviral response (Bellmann et al., 2019; Shelkovnikova et al., 2019) or nuclear pore defects (Lin et al., 2021), suggesting the contribution of environmental insults to FUS mislocalization. In addition, pathological mislocalization of FUS could also result from aggregation of other RNA-binding proteins, such as TAF-15 (Tetter et al., 2024) in FTLD-FET, or from pathological spreading of FUS from neighboring cells (Vazquez-Sanchez et al., 2024). Last, FUS mislocalisation could also result from increased genetic burden that ultimately drives this pathological hallmark in vulnerable neurons. The latter mechanism is supported by our observation of increased dementia-related genetic burden in ALSci patients that correlates with the extent of FUS regulon downregulation. It is however very likely that a conjunction of these different mechanisms leads to FUS mislocalisation and downregulation of the FUS regulon in vulnerable L5-ET neurons. Whatever the underlying cause of FUS mislocalization, our study demonstrates that its induction in adult CPNs is on its own sufficient to lead to ALS-ci relevant cognitive symptoms.

A second major result of our study is that FUS mislocalization drives genetic risk of cognitive and behavioral impairment in ALS. A previous survey of relationships between RBPs binding targets and ALS heritability showed that target binding sites of 6 RBPs, including TDP-43 and FUS, were associated with ALS risk(Megat et al., 2023). This finding is consistent with the major GWAS hit *UNC13A* being a TDP-43 target(Brown et al., 2022; Ma et al., 2022). However, gene targets of TDP-43 or FUS are cell-type dependent(Jeong et al., 2017), and it was critical to identify those in CPNs. Here we identify a set of genes regulated by FUS mislocalization in CPNs through a cross-species approach. This cross-species approach increases the robustness of target identification by conserving them across species and cell types, but is conservative. We cannot exclude that additional, human-specific FUS-regulated genes could be missing from the current FUS-regulon gene list. It is unlikely that all these FUS-regulated genes share a unique FUS-dependent regulatory mechanism as FUS is a multi-functional RNA-binding protein, involved at all steps of gene expression, from transcription to splicing, local translation or microRNA-mediated regulation (Lagier-Tourenne et al., 2010; Picchiarelli et al., 2019; Piol et al., 2023). It is also possible that some of these FUS regulon genes are indirect FUS targets and the precise mechanisms linking these genes to FUS will have to be determined on a gene-by-gene basis in further studies. Importantly, TDP-43 also regulates RNA levels of FUS-regulon genes suggesting that convergent loss of function of FUS and TDP-43 could synergize in leading to ALS neurotoxicity (Lagier-Tourenne et al., 2012).

ALS-linked genes were significantly enriched in the FUS regulon, and a substantial number of ALS genes were present in this regulon, including familial genes (*C9ORF72*, *KIF5A*, *ERBB4, FUS* itself), but also genes with common ALS-associated variants such as *NEK1*, *SCFD1* or *SARM1*. Interestingly, genes of the FUS regulon are associated with ALS with cognitive/behavioral impairment including *C9ORF72 (Diekstra et al., 2014), KIF5A* (Dulski et al., 2023; Saez-Atienzar et al., 2020), *FUS* (Aguzzoli et al., 2022; Broustal et al., 2010; Van Langenhove et al., 2010)*, ERBB4(Sun et al., 2020)*, or *SCFD1*(Chen et al., 2024). This finding suggests that the genes of the FUS regulon could be enriched in genes relevant for ALS-ci and ALS-bi rather than pure ALS. Consistently, we were able to uncover robust genetic associations between common variants in *FBXO16* and *WWOX* with ALS-bi and ALS-ci respectively. *WWOX* has been genetically associated with Alzheimer’s disease (Kunkle et al., 2019) and to risk of cognitive progression in Parkinson’s disease (Liu et al., 2021), and dementia in limbic-predominant age-related TDP-43 encephalopathy neuropathological change (LATE-NC), hippocampal sclerosis, and brain arteriolosclerosis (Dugan et al., 2022) indicating that this gene is broadly associated to cognitive deficits in multiple neurodegenerative diseases. *WWOX* encodes for a protein of poorly characterized function that has been suggested to indirectly modulate transcriptional activity and cell proliferation (Kosla et al., 2019). Its function in neurons is poorly documented, but it is worth noting that loss-of-function mutations are also associated with *WWOX*-related epileptic encephalopathy (WOREE) syndrome characterized by intractable epilepsy, severe developmental delay, motor deficits and premature death at the age of 2-4 years (Mallaret et al., 2014). Consistently, ubiquitous or neuronal deletion of *Wwox* in mice leads to a complex neurodevelopmental and neurodegenerative phenotype (Cheng et al., 2020; Repudi et al., 2021). Our results suggest that mild loss of *WWOX* could sensitize to cognitive impairment during ALS, thereby favoring an ALS-ci clinical picture.

We also show a genetic association between *FBXO16* and ALS-bi, and loss of FBXO16 in PTV carriers is sufficient to lead to multiple abnormalities in behavior, neuroanatomy and circulating biomarkers all consistent with fronto-temporal damage. *FBXO16* encodes for a largely uncharacterized F-box containing protein. F-box containing proteins are well known for their E3 ubiquitination ligase function in the SCF complex (SKP1-CULLIN1-F-box protein) and it is thus likely that FBXO16 is important in the ubiquitination and possibly degradation of specific, currently unknown, substrates(Wang et al., 2014). In cancer cells, FBXO16 has been shown to inhibit the beta-catenin pathway through proteosomal targeting of nuclear beta-catenin (Khan et al., 2019; Paul et al., 2019). Whether this function is related to behavioral impairment in *FBOX16* PTV carriers or in ALS-bi patients carrying *FBXO16* common variant remains to be determined. Nonetheless, our newly defined association between *FBXO16* and ALS-bi, indicates that perturbations in proteostasis likely contribute to disease pathogenesis.

In totality, our study demonstrates that additional FUS pathology is a contributor to cognitive impairment occurring in ALS patients and is related to the genetic risk of ALS-related cognitive and behavioral impairments. We further identify and replicate *FBXO16* as a novel gene contributing to behavioral impairments in ALS.

## Methods

### Patients tissue staining

#### Ethics

All clinical data including the ECAS were collected as part of Scottish Motor Neuron Disease Register (SMNDR) and Care Audit Research and Evaluation for Motor Neuron Disease (CARE-MND) platform (ethics approval from Scotland A Research Ethics Committee 10/MRE00/78 and 15/SS/0216) and all patients consented to the use of their data during life. All post-mortem tissues were collected via the Edinburgh Brain Bank (ethics approval from East of Scotland Research Ethics Service, 16/ES/0084) in line with the Human Tissue (Scotland) Act. Use of human tissue for post-mortem studies has been reviewed and approved by the Edinburgh Brain Bank ethics committee and the Academic and Clinical Central Office for Research and Development (ACCORD) medical research ethics committee (AMREC). BA39 was selected as a brain region that assesses language function and had been evaluated in at least 6 *C9ORF72* patients with 3 demonstrating impairment (as measured by clinically approved cut-offs measured by the Edinburgh Cognitive ALS Screening tool (Gregory et al., 2020). BA4 was used as a representative region of the motor cortex.

#### Immunohistochemical staining

Immunohistochemistry was performed on 4 µm-thick sections on superfrost charged slides. Sections were deparaffinized, followed by antigen retrieval using citric acid pH6. Sections were then stained using the Novolink Polymer detection system (DAB chromogenic staining) using FUS primary antibody (BioTechne NB100-595) at 1 in 500 dilution. Counterstaining was performed using lithium carbonate and haematoxylin. Slides were imaged using an EVOS M5000 microscope using brightfield settings at 10x and 40x magnification.

### Mouse breeding and genotyping

Mice were housed in our animal facility (Strasbourg), with a regular 12h light and dark cycle under constant conditions (21 ± 1°C, 60% humidity) and unrestricted access to food and water. Double transgenic mice were generated by crossing Thy1.2 CreERT2 mice (The Jackson Laboratory, JAX stock #012708 STOCK Tg(Thy1-cre/ERT2,-EYFP)HGfng/PyngJ) (Caroni, 1997; Feil et al., 1997; Young et al., 2008) with mice carrying the nuclear localization signal (NLS) of *Fus* flanked by two LoxP sequences (Mouse Clinical Institute of Strasbourg, Phenomin-ICS). We obtained offspring in which a tamoxifen-inducible Cre-mediated recombination aimed at deleting the NLS of FUS specifically in long projection neurons. We used mice containing two alleles with a LoxP-flanked Fus NLS sequence (FusLoxP/LoxP, referred to as “*Fus^exon15^*”) and homozygous double mutant mice (Thy1.2 CreERT2 FusLoxP/LoxP, referred to as “Thy1*-Fus^exon15^* mice”). Mice were carefully handled to reduce their stress and anxiety. Experimental protocols and animal care complied with the institutional guidelines (council directive 87/848, 19 October 1987, Ministry of Agriculture and Forestry, Veterinary Department of Health and Animal Protection) and international laws (directive 2010/63/UE609, 13 February 2013, European Community) and policies (personal authorization #I-67UnivLouis Pasteur-F1-04 for R.C.). The project has been approved by the ethics committee of Strasbourg (CREMEAS, reference number: FUS NLS inductible_#12173-2018062510569395).

### Primary neuronal cultures

Primary cortical cells were taken from homozygous *Fus^exon15^* E145 mouse embryos and cultured in complete neurobasal medium, i.e. Neurobasal medium (Thermofisher) supplemented with 1% Penicillin/Streptomycin (Sigma), 1%GlutaMAX (ThermoFisher) and 2% B27 (ThermoFisher). Dissected cortices were incubated with trypsin/EDTA (Sigma) and DNAse (50U/ml final Sigma), washed, and triturated to give a single cell suspension. Cells were cultured for 3-4 h in complex neurobasal medium complemented with 10% FBS (Thermofisher). From day 1 to day 14, we used complete neurobasal medium and changed the medium every 2-3 days. Cultured neurons were then exposed to an adenoviral vector (Ad-CRE-GFP, Vector biolab, #1700 and ad-CMV-GFP, Vector biolab, #1060 as control) at MOI 1, 10, and 100 either 5 or 14 days after plating.

### Induction of recombination in adult mice

The recombination was induced at 1.5-months of age by forced-feeding mice of both genotypes (*Fus^exon15^* and Thy1*-Fus^exon15^* mice) with 100 µL of tamoxifen at 20 mg/mL (Sigma) diluted in Corn oil (Sigma) for 5 consecutive days. We waited another month and a half before doing the different procedures (i.e. behavioral studies or dissection for biochemical analyses).

### Behavioral studies in mice

All behavioral studies were made in the morning during the light phase and the experimental procedure started after a 30-minute period of habituation to the experimental room. Mice were handled for at least five days to reduce stress and anxiety due to manipulation.

### Motor test

Mice were subjected to the inverted grid test to noninvasively assess the ability of mice to use sustained limb tension to oppose their gravitational force(Carlson et al., 2010). Each mouse was placed on a grid and was allowed to adapt to this environment for 15-20 s. The grid was then inverted and held approximately 35 cm over a cage containing 5 cm of wood chips. We assessed the hanging time, i.e. the time the mice hold before falling, for three trials with 5-minute breaks between each and a cut-off time of 5 min. The mouse body mass was measured shortly before the test to compute the minimal total sustained force that was exerted to oppose the gravitational force: holding impulse (N.s) = averaged hanging time (s) × body mass (g) × 0.00980665 (N.g^−1^).

### Nest building test

Nesting abilities were measured based on a previously described protocol(Deacon, 2006). One hour before the dark phase, enrichment items were removed from individual home cages and 3 g of pressed cotton pieces (i.e. approximately 6 pieces) were given to each mouse. After 24h nests were assessed with the five-point scale used by Deacon(Deacon, 2006). Untorn cotton pieces were counted and weighed.

### Three-chamber social task

The experimental procedure was adapted from Gascon E et al(Gascon et al., 2014). The three-chamber box (59 × 39.5 × 21.5 cm) is made of transparent Plexiglas (Noldus Information Technology, Wageningen, The Netherlands) and is divided into three chambers (one middle and two side chambers) of equal size (18.5 × 39.5 cm) by the walls with a square opening (7 × 7 cm) that could be closed by a slide door. Each tested mouse was placed in the middle of the central compartment that was kept empty while the two others compartments contained an openwork cylindrical cage (20 × 10 cm diameter made of transparent Plexiglas bars). After the habituation session (one trial with free access of the arena containing two empty cages), mice were placed three times (15-minute inter-trial interval) in the arena containing a cage with a mouse (referred to as “mouse””) and an empty cage. Mice were placed one last time in the arena containing a cage with the known mouse (i.e. the mouse already meet tree times during the previous phase) and a cage with a novel mouse (referred to as ““Novel mouse”). For all sessions, mice were free to explore the arena for 5 min while being recorded. For all sessions, mice were free to explore the arena for 5 min while being recorded. Two parameters were measured using the EthoVision XT software (Noldus Information Technology) : total distance travelled and velocity. The time spent contacting the empty cage and the strangers was measured by hand. The recognition index (RI) was calculated according to this formula: RI = contact time with stranger 2 / total contact time × 100.

### Resident-intruder social task (social interaction in home cage)

Briefly, after 30 minutes of habituation to the test room resident mouse was allowed to freely roam in its home cage without the cage top for 1 min. A novel male intruder mouse (non-littermate of the same background, same age, and similar weight) was introduced in the opposite corner as the resident. We recorded the interaction for 5 min to assess social activities: total physical interaction (active exploration of the intruder) comprising investigating, grooming, following and sniffing(Scekic-Zahirovic et al., 2021).

#### Dark/light box test

The apparatus consisted of a polypropylene cage (44 × 21 × 21 cm) separated into two compartments with a small opening (12 × 5 cm) at floor level. The first compartment (28 cm long) was open-topped, transparent, and brightly illuminated by white light from a 40-watt desk lamp positioned above the lit chamber (140 lux). The second compartment is smaller (14 cm long) and covered at the top with black Plexiglas and painted black on all sides. Mice were individually placed in the center of the dark compartment and allowed to freely explore the apparatus for 5 min. The apparatus was cleaned with a 30% ethanol solution between subjects. We assessed the number of light/dark transitions between the two compartments, the latency to first get out of the dark box, and the total time spent in the light compartment.

#### Object recognition test

The novel object recognition test was performed to investigate learning and memory in mice(Lueptow, 2017). Each mouse was placed in the middle of an open and squared arena containing two objects. The position of these objects was the same for all sessions and the objects and arena were cleaned with 70% (v/v) ethanol. For the habituation (day 1), mice were placed two times, with a 10-minute break in between, in the arena containing two identical objects (empty tips boxes). Object recognition was tested after a short retention time (10 min) to evaluate short-term memory (day 2) and after a longer one (24h) to look at long-term memory (days 3 and 4). Mice were first placed in the arena with two identical objects (either two water glasses for short term memory or two glass bottles for long-term memory). Then, for memory assessment, one of the previous objects was replaced by a novel one (a 15-cm high vial for short-term memory and stemmed glass for long term memory). For all sessions mice were free to explore the arena during 10 min while being recorded. Two parameters were measured using the EthoVision XT software (Noldus Information Technology): total distance travelled and velocity. The times spent exploring the identical objects, the known object (Tk) or the novel one (Tn) were manually determined. The recognition index (RI) was calculated according to this formula: RI = exploration time of the novel object (Tn) / total exploration time (Tk + Tn) × 100.

#### Sucrose splash test

The protocol, adapted from Isingrini et al. (2010),(Isingrini et al., 2010) consists of spraying a 10% sucrose solution on the dorsal coat of a mouse in its home cage. Due to sucrose’s viscosity, the sucrose solution dirties the mouse fur and animals initiate grooming behaviors. Once the sucrose solution was applied, latency (time between spray and initiation of grooming) and duration of grooming were recorded for 5 minutes.

#### Sucrose preference test

The Sucrose Preference Test (SPT) is a reward-based test used as an indicator of anhedonia. We used a protocol adapted from Serchov et al.(Serchov et al., 2016). Briefly, mice were presented for three days in their home cage with two identical-looking bottles, but one contains water while the other contains a sucrose solution (1%, EMD Millipore, catalog number: 573113). Consumption of water and sucrose solution was measured daily, and the positions of the two bottles were interchanged each day to reduce any risk of lateral bias. Sucrose preference (SP) was calculated as a percentage of the volume of sucrose intake over the total volume of fluid intake and averaged over the three days of testing according to this formula: SP = V(sucrose solution)/[V(sucrose solution)+V(water)] x 100%.

### Electrocorticography in mice

#### Surgery

Surgery procedures were similar to previous studies(Guillot et al., 2025; Scekic-Zahirovic et al., 2024). Briefly, mice were deeply anaesthesized using 4% isoflurane (Baxter). Prior to the procedure, injectable meloxicam (Metacam, 10mg/mL, Boehringer Ingelheim) was provided subcutaneously to the animals. After local subcutaneous injection of lidocaine (Lurocaine, 2mg/kg, Vetoquinol), animals were stereotaxically implanted with electrodes at the following coordinates: Reference electrode: Bregma +0.50mm, midline +0.00mm; Ground electrode: Bregma −3.00mm, midline −3.00mm; Recording electrode 1: Bregma +2.50mm, midline +2.00mm; Recording electrode 2: Bregma −2.50mm, midline −2.00mm. The incision was closed using dental cement (Super-Bond C&B). The animals were returned to their home cage and given meloxicam (Metacam, 5mg/mL, Boehringer Ingelheim) *ad libitum* in their drinking water for five days after the procedure.

#### Analysis

Electrocorticography (ECoG) recordings were conducted on Thy1^CRE-ERT2^/*Fus^exon15^* mice and their respective control littermates at 3 and 5 months of age, over a 24-hour period. ECoG was recorded using LabChart (LabChart version 5, ADInstruments, USA). ECoG signals were collected with a 2,000Hz sampling rate. The Python package EEVop(Guillot, 2023) was used to analyze the ECoG traces. To remove the powerline (50Hz) and its harmonics up to 200Hz, ECoGs were notch-filtered. A Z-score was used to normalize each ECoG recording. The Z-score of each recording was compared with the mean Z-score of the control animals. Only events that exceeded the baseline Z-score were counted. To ensure proper removal of background noise, a window discriminator of four times the baseline standard deviation was used for event detection.

### Immunostaining in mice and imaging

Immunolabelling experiments were performed 1.5 months after tamoxifen administration. The mice were deeply anaesthetized using an intraperitoneal injection of pentobarbital (60 mg/kg) and then transcardially perfused with cold 4% PFA in 0.01 M phosphate-buffered saline (PBS). After dissection (brain and spinal were splitted), brains were post-fixed for 24 hours and then included in agar 4% and serial cuts of 40 µm thick were made using vibratome (Leica Biosystems, S2000). After three washes (0.1M PBS), tissues were incubated for a 30 min period in citrate sodium buffer (pH 6, 10 mM) at 85°C and then left for 90 min in PBS with 0,5% Triton X-100 and 5% donkey serum under slow agitation. The primary antibodies were incubated in 0.1% Triton X-100 and 1% donkey serum in PBS for 24h at 4°C: rabbit@FUS N-ter (Protein Tech, #11570-1-P, 1/250), rabbit@FUS C-ter (Bethyl, #A300-294, 1/500), chicken@GFP (Abcam, #a13970, 1/1000), goat@ChAT (Millipore, #AB114P, 1/1000), mouse@NeuN (Millipore, #MAB377; 1/250), goat@Iba1 (abcam, #ab5076, 1/250), rabbit@GFAP (Dakocytomation, 1/250). After three washes, the secondary antibodies were incubated in 0.1% Triton X-100 in PBS overnight at 4°C: Alexa Fluor 488 donkey anti-chicken IgG H&L (JacksonLab, #703-545-155 5, 1/500); Alexa Fluor 647 donkey anti-rabbit IgG H&L (JacksonLab, #711-605-152, 1/500); Alexa Fluor 555 donkey anti-mouse IgG H&L (Molecular Probe, #A21203, 1/500), Alexa Fluor 555 donkey anti-goat IgG H&L (JacksonLab, #705-605-147, 1/500). A chromatin staining with DAPI was performed between the two final washes. The sections were oriented and mounted in Mowiol 4-88 medium (Sigma).

Image acquisition was performed at the Imaging Core Facility of Strasbourg Biomedical Research Centre (University of Strasbourg, INSERM, PIC-STRA, UMS 38). Fluorescence images were acquired using a Zeiss (LSM 710 NLO) confocal microscope with a 20× Zeiss Plan Apochromat oil-immersion lens (NA 1.40) controlled with the Zen software (ZEN 2012 Black edition, Carl Zeiss Microscopy GmbH, Germany). Images were acquired to a resolution of 2048×2048 pixels at subsaturating laser intensities for each channel. To evaluate the percentage of recombined neurons (in the brain) and motoneurons (in the spinal cord), we performed manual counting with FIJI free software(Schindelin et al., 2012). The measurement of the number of neurons in the frontal and motor cortex was made using a homemade macro.

### Western blotting and subcellular fractionation

#### Subcellular fractionation and Western Blotting

Tissues were washed in 1x PBS and lysed in NE-PERNuclear and Cytoplasmic Extraction (Thermo Scientific, 78835) according to the manufacturer’s instructions. Protein extracts were dosed by BCA Assay (Interchim, UP95424A). Proteins were denatured and SDS page was performed with 30 μg of cytoplasmic proteins and 10 μg of nuclear proteins on Criterion TGX stain-free gel 4–20 % (Biorad, 5678094). Proteins were blotted on nitrocellulose membrane using semi-dry Transblot Turbo system (BioRad, France) and blocked with 10 % non-fat milk during 1 h. Primary antibodies (Rabbit anti-FUS C-ter targeting the NLS (Bethyl, #A300-294 A, 1/2000), Rabbit anti-FUS N-Ter (Bethyl, #A-300-293 A, 1/2000), Sheep anti-SOD1 (Calbiochem, 574597, 1/1000), Rabbit anti-HDAC1 (Bethyl, #A300-713 A, 1/1000) were incubated overnight at 4 °C in 3 % non-fat milk. Washing was proceeded with washing buffer (Tris pH 7.4 1 M, NaCl 5 M, Tween 20 100 %) and secondary antibodies (anti-rabbit HRP (PARIS, #BI2407,1/5000), anti-sheep HRP (Jackson, #713-035-147, 1/5000); anti-Rat HRP (Rockland, 612-1102, 1/5000) were incubated for 1h30 at room temperature. After successive washes, proteins were visualized with chemiluminescence using ECL Lumina Forte (Millipore, France) and chemiluminescence detector (Bio-Rad, France). Total proteins were detected with stain-free gel capacity (Biorad, 5678094) and used to normalize for protein loading.

### RT-qPCR

#### Genomic DNA isolation and PCR

Genomic DNA was extracted from hippocampi and lumbar spinal cord using KTT Buffer (Tris HCl, Triton and KCL) with Proteinase K (Sigma, 20mg/mg). DNA was purified using Phenol/Chloroforme (Sigma, P2069) and washed with 70% Ethanol. Polymerase chain reaction (PCR) was performed with MasterMix Taq DNApolymerase (VWR International, Ref. 733–1320) and the following primers : LF2 : 5’-CTA-GCC-CAC-CTC-CTA-ATC-CTC-ATC-AC-3’; LR2 : 5’-CCA-CAC-TTT-AGG-TTA-GTC-ACA-GAT-CAG-C-3’ and LXR : 5’-GGA-TCC-GCG-GGA-AGT-TCC-TAT-AC-3’. PCR1 correspond to LF2 + LR2 and PCR2 correspond to LF2 + LXR. The following program was used : 5 min 95°C; (30 s 95°C; 30 s 62°C; 30 s 72°C) x 34; 5 min 72°C). 10 μL of the PCR products were loaded on a 2 % agarose (Euromedex, Ref.D5-E) gel electrophoresis with Low Molecular Weight DNA Ladder (NEB, Ref. N3233L) and stained with ethidium bromide using standard procedures.

#### RNA extraction and quantitative PCR

Total RNA was extracted from hippocampi and lumbar spinal cord using TRIzol® reagent (Life Technologies). 1 μg of RNA was reverse transcribed with iScript™ reverse transcription (Biorad, 1,708,841). Quantitative PCR was performed using Sso Advanced Universal SYBR Green Supermix (Bio-Rad 1,725,274) and quantified with Bio-Rad software. Gene expression was normalized by calculating a normalization factor using actin, TBP and pol2 genes according to GeNorm software. Primer sequences are provided in Table XX.

#### RT-PCR

1 μg of RNA was reverse transcribed with iScript™ reverse transcription (Biorad, 1,708,841). PCR was performed using in 25 μL microtubes with MasterMix Taq DNApolymerase (VWR International, Ref. 733–1320) and the following programs: Intron 6 retention (5 min 95 °C, (30 s 95 °C, 30 s 56 °C, 30 s 68 °C) x 30; 5 min 68 °C), Intron 7 retention (5 min 95 °C, (30 s 95 °C, 30 s 61 °C, 30 s 68 °C)x 30; 5 min 68 °C), 10 μL of the PCR products were loaded on a 2 % agarose (Euromedex, Ref.D5-E) gel electrophoresis with Low Molecular Weight DNA Ladder (NEB, Ref. N3233L) and stained with ethidium bromide using standard procedures. The signal intensity of each of the two bands was quantified individually, and a percentage of intron retention was computed according to this formula: (intensity of Intron + band) / (intensity of Intron + band + intensity of Intron - band) × 100.

### Single nuclei RNA sequencing

#### 10x Genomics Chromium single-cell 3′ library construction

Cell capture, amplification and library construction on the 10x Genomics Chromium platform were performed according to the manufacturer’s publicly available protocol. Briefly, viability was assessed by trypan blue exclusion assay, and cell density was adjusted to 175 cells per microliter. In total, 7,000 cells were then loaded onto a single channel of a 10x Chromium chip for each sample. The 10x Genomics Chromium technology enables 3′ digital gene expression profiling of thousands of cells from a single sample by separately indexing each cell’s transcriptome. Primers containing (i) an Illumina R1 sequence (read 1 sequencing primer), (ii) a 16-nucleotide 10x Barcode, (iii) a 10-nucleotide UMI, and (iv) a poly-dT primer sequence were released and mixed with cell lysate and Master Mix. Incubation of the GEMs then produced barcoded, full-length cDNA from polyadenylated mRNA. After incubation, the GEMs were broken and the pooled fractions were recovered. Full-length, barcoded cDNA was then amplified by PCR to generate sufficient mass for library construction. Enzymatic fragmentation and size selection were used to optimize the cDNA amplicon size before library construction. R1 (read 1 primer sequence) was added to the molecules during GEM incubation. P5, P7, a sample index and R2 (read 2 primer sequence) were added during library construction via end repair, A-tailing, adaptor ligation and PCR. The final libraries contained the P5 and P7 primers used in Illumina bridge amplification. The described protocol produced Illumina-ready sequencing libraries. A single-cell 3′ library comprises standard Illumina paired-end constructs that begin and end with P5 and P7. The single-cell 3′ 16-bp 10x Barcode and 10-bp UMI are encoded in read 1, while read 2 is used to sequence the cDNA fragment. Sample index sequences were incorporated as the i7 index read. Read 1 and read 2 are standard Illumina sequencing primer sites used in paired-end sequencing. Sequencing the library produced a standard Illumina BCL data output folder. The BCL data include the paired-end read 1 (containing the 16-bp 10x Barcode and 10-bp UMI) and read 2 as well as the sample index in the i7 index read.

#### Processing of *Fus^Δ^*^NLS^ mice single-cell RNA-seq data

Raw FASTQ files were aligned to the pre-mRNA annotated mouse reference genome GRCm10. Cell Ranger v.4.0 with default parameters was used to demultiplex and align our barcoded reads with the Ensembl transcriptome annotation (GRCm10). The majority of our downstream analysis was conducted using the R programming language (v.4.0.5 for harmonization and clustering, v.4.1.0 for annotation and downstream visualization) and the RStudio integrated development environment. A recent report suggested that filtering cells with greater than 10% mitochondrial reads is the preferred baseline for human tissue, and that for brain tissue a higher threshold may even be optimal. Thus, a mitochondrial percentage that was the higher of either 10% of reads or the two absolute deviations above the median for mitochondrial reads within the sample was chosen as a threshold. Cells below this threshold with between 500 and 10,000 UMIs were retained for downstream analysis. All ribosomal genes, mitochondrial genes and pseudogenes were removed, as they interfered with the downstream differential gene expression.

#### Processing of ALS patients’ single nuclei RNA sequencing

Raw FASTQ files were downloaded from Pineda et al.(Pineda et al., 2024) and aligned to the pre-mRNA annotated human reference genome GRCh38 using Cell Ranger v4.0 (10x Genomics, Pleasanton CA). Introns were annotated as ‘mRNA’, and intronic reads were included to quantify expression. Quality control criteria were used as previously described (Bakken et al., 2021) so that for Cv3, criteria were: more than 500 (non-neuronal nuclei) or more than 1,000 (neuronal nuclei) genes were detected and doublet score was less than 0.3.

#### Clustering

Nuclei were grouped into transcriptomic cell types using an iterative clustering procedure. Read counts were summed, and log_2_-transformed expression was centered and scaled across nuclei. The graph-based clustering approach implemented in Seurat (v.5) was used to cluster our cells. In brief, a *k*-nearest neighbors graph based on Euclidean distance in our corrected mNN space was calculated and used to derive refined edge weights based on Jaccard similarity. The Louvain algorithm was then applied to iteratively delineate a population structure on our dataset. This was implemented with the FindNeighbors and FindClusters functions in Seurat. A UMAP projection of our dataset was computed with the RunUMAP function for visualization (Figure 1). Differentially expressed genes (DEGs) for a given cluster were identified by using Seurat’s FindAllMarkers function with a Wilcox test and comparing each cluster with every other cluster under the same subclass, with logfc.threshold set to 0.25 and min.pct set to 0.5. The union of up to 50 genes per cluster with the highest average logFC was used. The average log_2_ expression of the DEGs was then used as input for the build_dend function from scrattch.hicat to create the dendrograms.

##### Cell type annotation

A curated set of known major cell type markers based on Bakken et al., 2021 was used to annotate individual cells with their expected cell type and assign a confidence score to each annotation. For ambiguous subpopulations, we assigned identity based on visually distinct, well-defined gene expression domains reproducibly identified by Leiden clustering at multiple resolutions. Descriptive labels were determined by the unique co-expression of marker genes identified by Seurat for each subclass.

#### Pseudobulk differential expression analysis

Cell type-specific pseudo-bulk differential gene expression (DGE) groups were built based on hierarchical clustering and Euclidean distance between each cell subclass. The pseudobulk methods employed the edgeR for analysis of aggregated read counts. Briefly, for cells of a given type, we first aggregated reads across biological replicates, transforming a genes-by-cells matrix to a genes-by-replicates matrix using matrix multiplication. For each differential expression cluster, only genes present in at least 10% of cells were retained for analysis. Counts were depth normalized, scaled by library-size, and log_2_-transformed. Pseudo-bulk expression profiles were computed by averaging normalized log-counts within each cluster for each unique sample. We used surrogate variable analysis(Leek et al., 2012) (implemented in the SVA R package) to identify and remove sources of unknown variance. We used the quasi-likelihood F-test approach (edgeR-QLF) and gene were considered differentially expressed for FDR<0.05.

#### Cross-species consensus co-expression analysis

To facilitate comparison across species, mouse gene identifiers were re-annotated with human Ensembl gene orthologs using biomaRt, an R interface with the Biomart database (www.biomart.org). Only identifiers that were common to both human and mouse meta-sets were retained. A consensus network represents a single network arising from multiple sources of data constructed from the weighted average of correlation matrices from both the human and mouse in this study. By definition, consensus modules are the branches of a clustering tree developed from a consensus gene dissimilarity, comparable to the single-network approach; consensus modules contain genes that are closely related in both networks, i.e., the modules are present in both networks. After scaling the network (consensus scaling quantile = 0.2), a threshold power of 14 was chosen (as the smallest threshold resulting in a scale-free R^2^ fit of 0.9). The consensus tree was then built which identified 4 modules for which correlation to phenotype (ALS) and adjusted-pvalue (FDR) were calculated. This approach led to the identification of a final set of 4 modules.

#### Violin plot of eigengene expression

For each module and corresponding cell type, the module eigengene was calculated in control and ALS conditions using the moduleEigengenes() function. Comparison across conditions was calculated using a Wilcoxon rank-sum test and considered significant when Bonferroni corrected p-value < 0.05.

### ENCODE RNA-binding proteins (RBP) knock-down experiments

A list of 217 RBPs and their counts files were downloaded from the ENCODE consortium (https://www.encodeproject.org/). Counts files for each RBPs were downloaded and normalized to library size and internal controls. A final list of 217 RBPs was used to calculate the eigengene expression of the FUS regulon. Each RBPs was then ranked according to the magnitude of the FUS regulon dysregulation.

### Olink proteogenomics study cohort

Olink proteomic profiling was conducted on blood plasma samples collected from 54,967 UK Biobank participants using the Olink Explore 3072 platform. This platform measured 2,923 protein analytes, reflecting 2,941 unique proteins measured across the Olink panels that comprise the 3072 panel. For whole-exome sequencing-based proteogenomic analyses, we analyzed the 52,217 samples with available paired-exome sequence data. We identified the participants with European genetic ancestry based on HapMap projection. We performed finer-scale ancestry pruning of these individuals, retaining those within 4 s.d. from the mean across the first four principal components, resulting in a final cohort of 46,327 individuals for the proteogenomic analyses. Finally, we built metadata on neurodegenerative diseases diagnosed after enrollment (AD, PD, ALS, VaD, AD) and carriers’ status of *FBXO16* PTV (*FBXO16*). We filtered individuals to have age-and sex matched controls for each disease resulting in a final cohort of 24,557 individuals. To ensures a minimum number of observations per group, proteins with a high fraction of missing values (>75% of participants) were excluded from the regression analysis. In addition, groupwise outlier removal of protein measurements (±1.5 × IQR) was performed before regression analysis because the presence of outliers can severely affect the resulting test statistics, potentially increasing the rate of false negatives in the initial biomarker discovery phase. Linear regression adjusted for age and sex, was performed for each protein and disease. Genome-wide significant association was set at Bonferroni corrected p-value < 0.05 / 2,941.

### Brain MRI UK Biobank data acquisition

UK Biobank <u>brain</u> imaging has been described as a resource (Miller et al., 2016), where the reader can find a number of examples describing how it can be accessed and used for research. Three dedicated imaging centers are equipped with identical scanners (3T Siemens Skyra, software VD13) for brain imaging scanning using the standard Siemens 32-channel receive head coil. The brain MRI protocol is performed using a 3 Tesla Siemens Skyra scanner (Siemens Healthineers, Erlangen, Germany) with VD13 software and a 32-channel head coil. The protocol includes three structural MRI scans; T1, T2 fluid attenuation inversion recovery (FLAIR) and susceptibility-weighted MRI (swMRI), as well as diffusion MRI (dMRI) and resting and task functional MRI (fMRI). T1 scans allow precise volumetric measures of the whole brain, as well as specific cortical and subcortical regions. An automated processing pipeline for brain image analysis and quality control was established for UKB at the University of Oxford’s Wellcome Centre for Integrative Neuroimaging (WIN/FMRIB). This pipeline is primarily based around FSL (FMRIB’s Software Library), and other packages such as FreeSurfer. When acquired at the imaging center, the images are reconstructed from *k*-space on the scanner computer and saved initially as DICOM files. The processing pipeline then converts these files to the NIFTI format and undertakes pre-processing (e.g., correcting for head motion and other artefacts) as well as automated quality control that identifies issue with the equipment (e.g., coil failure) and artefacts specific to the participant or scanning session (e.g., excessive head movement). The NIFTI files for the T1 are provided to researchers, as these are suitably “defaced” to remove the possibility of re-identification of any individual participant. The pipeline also automatically generates thousands of IDPs, such as regional grey matter volume from T1 scans, fractional anisotropy, axial diffusivity and mean diffusivity measures from dMRI scans. These IDPs have been made available to researchers with authorized access and were used for GWAS association with FBXO16 carriers’ status.

### Grey matter volume, area and thickness analysis in *FBXO16* PTV carriers

Grey matter volumes, area and thickness were produced using the *recon-all* script, in cross-sectional mode, of FreeSurfer version 5.3 (Fischl, 2012) on each T1-weighted scan. The standard pre-processing steps of the FreeSurfer image analysis suite were first implemented, including removal of non-brain tissue, segmentation of the subcortical white matter and deep grey matter structures, intensity normalization, tessellation of the grey matter–white matter boundary, and automated topology correction. Following quality control steps for segmentation accuracy, overall volumes of cortical structures and total intracranial volume estimates (eTIV) were retrieved from each subject. A dual pipeline was implemented exploring cortical thickness alterations and morphometric changes using voxel-based morphometry (VBM). Following pre-processing and cortical segmentation in FreeSurfer, average cortical thickness, volume and area values have been retrieved from 34 cortical regions in each hemisphere separately as per the Desikan–Killiany atlas(Desikan et al., 2006). Cortical thickness values in the following lobes and corresponding subregions regions were appraised, Frontal lobe (13 ROIs): Superior Frontal, Rostral and Caudal Middle Frontal, Pars Opercularis, Pars Triangularis, and Pars Orbitalis, Lateral and Medial Orbitofrontal, Precentral, Paracentral, Frontal Pole, Rostral Anterior cingulate, Caudal Anterior cingulate, Parietal lobe (7 ROIs): Superior Parietal, Inferior Parietal, Supramarginal, Postcentral, Precuneus, Posterior cingulate, cingulate isthmus, Temporal lobe (9 ROIs): Superior, Middle, and Inferior Temporal, Banks of the Superior Temporal Sulcus, Fusiform, Transverse Temporal, Entorhinal, Temporal Pole, Parahippocampal, Occipital lobe (4 ROIs): lateral Occipital, Lingual, Cuneus, Pericalcarine, and the Insula (1 ROI). In addition to cortical thickness analyses, voxel-based morphometry was also performed to evaluate anatomical patterns of signal intensity reductions in mutation carriers. Regional effects of FBXO16 PTV status on cortical thickness, area and volume were tested by general linear models (GLM) across the entire cortex controlling for age and sex. Permutation-based non-parametric inference and the threshold-free cluster enhancement (TFCE) approach were used to test for differences between study groups and significance was set at FDR < 0.05

### Connectometry

For DTI preprocessing of brain MRI, the DSI Studio software (version May 2024, http://dsi-studio.labsolver.org) was utilized. This program, equipped with open-source images, addressed eddy current and phase distortion artefacts, generated a mask through thresholding, smoothing, and defragmentation, and underwent a quality control step for DTI. Subsequently, diffusion data were reconstructed using the DTI method and generalized q-sampling imaging (GQI) with a diffusion sampling length ratio of 1.25 (Yeh et al., 2010). The connectometry analysis incorporated DTI-based metrics, including FA, MD, AD, and RD values, and GQI-based QA values, extracted as the local connectome fingerprint (Yeh et al., 2016) When correlated with the apnea-hypopnea index (AHI), a representative measure of OSA severity, diffusion MRI connectometry demonstrated changes in FA, MD, AD, RD, and QA values. Non-parametric Spearman partial correlation, removing the effects of sex and age via a multiple regression model, was used to derive these correlations. A T-score threshold of 2.5 guided deterministic fiber tracking for correlational tractography.

### LD-score regression and partitioned heritability for ALS

We used the summary association statistics from the latest GWAS of ALS (van Rheenen et al., 2021). FUS regulon genes were mapped on single nuclei ATACseq data on major brain cell types (Corces et al., 2020) leading to 17,280 cell type-specific ATAC peaks associated with the FUS regulon (n=738 genes). As a positive control, we used the latest ALS GWAS and extracted SNP with suggestive association with ALS (p<5.10^−06^). Each SNP were mapped to the snATACseq using the same procedure leading to 13,278 ATAC peaks used as input to ALS-GWAS LDSC analysis. As a negative control, we randomly selected 15,000 ATAC peaks in a pool of 252,276 non-overlapping peaks across 6 major brain cell types. For each of the above-created sets, we ran S-LDSC (Finucane et al., 2018) to generate the LD score of each variant in each annotation using the same procedure. Regression SNPs, that are used by S-LDSC to estimate *τ* from marginal association statistics, were obtained from the HapMap Project phase (International HapMap, 2003). Using the LD score for each annotation and the marginal statistics obtained from the trait phenotypes (GWAS-ALS), we computed the enrichment and calculated the p-value for each annotation(Finucane et al., 2018).

### GWAS of ALS-bi/ci in the AnswerALS cohort

Whole genome sequencing from 631 ALS patients from the AnswerALS cohorts used were preprocessed for GWAS analysis (Baxi et al., 2022). Duplicate individuals were removed (king-cutoff = 0.084). Population structure was assessed by projecting 1000 G principal components (PCs) and outliers from the European ancestry population were removed (> 4 SD on PC1-4). In total, 546 ALS cases passed quality check analysis. WGS SNPs variants were mapped to genes using the SNP-to-Gene approach as described in MAGMA(de Leeuw et al., 2015) leading to a final set of 143,188 variants used for GWAS analysis. After quality control, a null logistic mixed model was fitted using PLINK with principal component (PC)1–PC10, sex, age at onset and APOE genotype as covariates. To assess any residual confounding due to population stratification and artificial structure in the data, we calculated the LDSC intercept using SNP LD scores calculated in the HapMap3 CEU population. Genome-wide significance was set at 5.10^−07^.

### Statistics

Values from each animal were averaged for each genotype group and data are expressed as mean ± standard error of the mean (SEM). Statistical analysis was performed using the GraphPad Prism 8 software with a significant threshold set at p < 0.05. Analysis of variance (ANOVA) was used because it was justified by the independence and normality of results and the equality of variances. Details of the analyses are systematically given in the figure legends. Normality and homoscedasticity were both assessed prior to analysis using Shapiro-Wilk and Bartlett’s test, respectively. Statistical analysis of two groups were performed using independent Student’s t-tests with Statsmodels package

## Data Availability

All data produced in the present study are available upon reasonable request to the authors

## Resource availability

All snRNAseq datasets have been deposited on GEO.

*Fus*^exon15^ mice are available upon request to the corresponding authors.

## Acknowledgements

The authors would like to thank the technicians of the animal facilities, namely Célian Pitard, Christiane Rieger and Claudia Spielmann, for their care of the mice. We are grateful to PICSTRA Imaging platform (UMS38, CRBS) and PEFRE animal facility platform (UMS38, CRBS for technical assistance and support. We acknowledge all persons involved in the mouse generation project at the ICS (Strasbourg, France) for their assistance with project design and data analysis. This work was supported in part by the Intramural Research Program of the National Institutes of Health, the National Institute on Aging (1ZIAAG000933). This work was funded by Agence Nationale de la Recherche (ANR-16-CE92-0031, ANR-19-CE17-0016, ANR-20-CE17-0008, ANR-21-CE17-0039, ANR-24-CE37-4064), by the Interdisciplinary Thematic Institute NeuroStra, as part of the ITI 2021-2028 (Idex Unistra ANR-10-IDEX-0002, ANR-20-SFRI-0012) by Fondation Bettencourt (Coup d’élan 2019 to LD), Fondation pour la recherche médicale (FRM, DEQ20180339179), Axa Research Funds (rare diseases award 2019, to LD), Association Francaise de Recherche sur la sclérose latérale amyotrophique, the Association Française contre les Myopathies (AFM-Téléthon, #23646, #28944)). LD is USIAS fellow 2019).

## Author contributions

Mouse breeding genotyping were performed by C.D.T., S.D. and R.C. Mouse tissues were processed by R.C., F.L. and A.B. Histological analysis in mice was conducted by R.C., F.L., A.B, and P.K. The *Fus* mouse model generation was conducted by M-C.B. Behavioral tests were conducted by R.C., F.L., A.B, and C.D, while the analysis was performed by R.C and S.D. EEG surgery and analysis were performed by S.J.G. and G.S.L. Single-nucleus RNA sequencing analysis, cross-species integration, genetic analyses, and UK Biobank data mining were conducted by S.M. Human Pathology was performed by F.M.W. and J.M.G.. SnRNAseq of ALS and ALS-ci patients was performed and analyzed by S.M., C.G.R., C.A.J. and H.P.. B.T., A.C., R.R. and J.S. contributed with clinical and genetic data as well as research tools. Funding was secured by R.C. and L.D. The work was conceptualized, coordinated and supervised by R.C., LD and SM. The manuscript was written by R.C, L.D. and S.M.

## Supplemental information

Document with Figures S1–S8. Tables S1 & S2.

