## Supplementary material for "FUS Mislocalization Rewires a Cortical Gene Network to Drive Cognitive and Behavioral Impairment in ALS": Figures S1-S8

**CASSEL and collaborators.**

### Supplementary figures

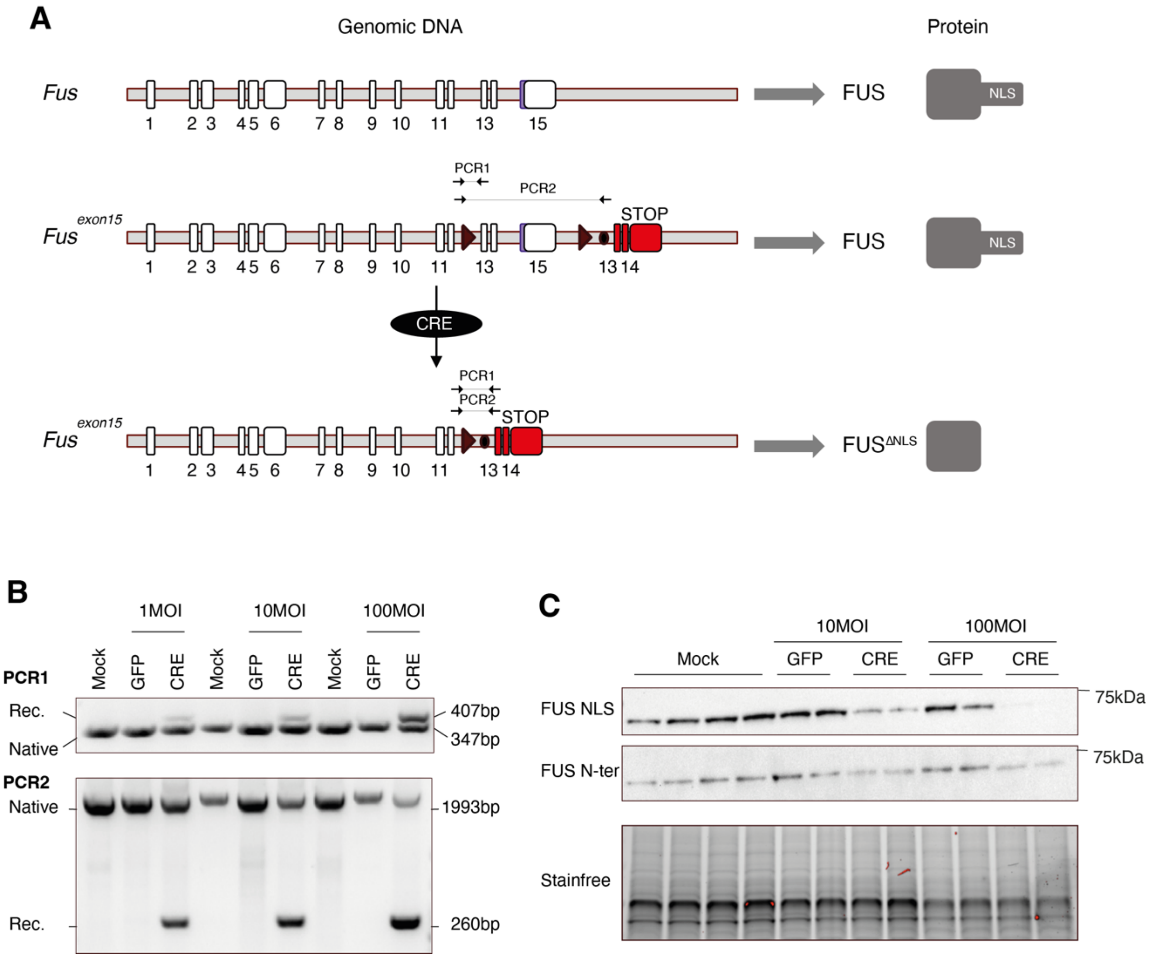


**Figure S1: design and *in vitro* validation of *Fus*$^{exon15}$ allele.**

- **A.** Schematic representation of the mouse wild-type *Fus* gene locus (upper scheme), with encoded protein (right). The middle scheme depicts the engineered *Fus*$^{exon15}$ allele, with the location of PCR primers for genotyping, and the lower panel shows this allele after CRE recombination, leading to a truncated FUS protein. Inserted cassettes are indicated in red, the nuclear localization signal (NLS)in blue and loxP sites are black triangles.
- **B.** Primary cortical neurons were transduced with the indicated MOI of Ad-CRE, Ad-GFP or mock. DNA was extracted and PCR was performed using primers shown in A.
- **C.** Immunoblot analysis of FUS protein in neurons treated as in B using a combination of two different antibodies targeting either the NLS or the N-terminal part of FUS. Molecular weight markers are shown on the left, and apparent molecular weight is indicated on the right. Stain-free image is provided as a loading control.

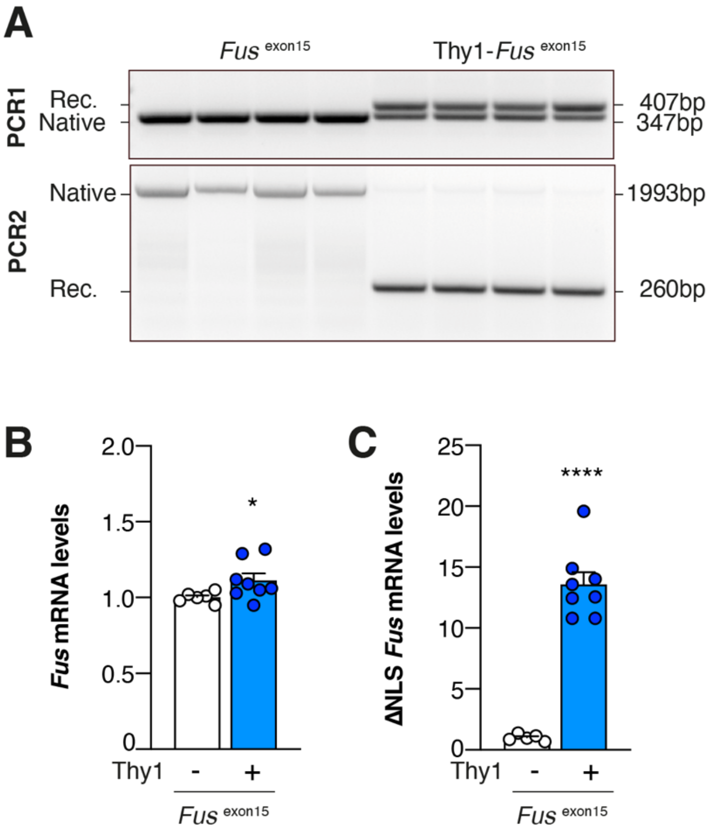


**Figure S2: Additional *in vivo* validation of *Fus* recombination and expression**

**(A)** Representative genotyping results obtained by PCR from the cerebral cortex of Thy1$^{CRE-ERT2}$/ *Fus*$^{exon15}$ mice and control mice showing recombination of the *Fus* locus as expected (260 bp vs 347 bp for the wild-type locus).

**(B-C)** RT-qPCR results of total *Fus* mRNA expression (B) and expression of the ΔNLS mRNA (after recombination, C) in the cortex of of Thy1$^{CRE-ERT2}$/ *Fus*$^{exon15}$ mice and control mice.

Results are expressed as mean ± SEM. Statistical analyses are performed with two-tailed Mann Whitney test: **** $p < 0.0001$, * $p < 0.05$, n = 6 for control mice (in black), n = 8 for Thy1$^{CRE-ERT2}$/ *Fus*$^{exon15}$ (in blue).

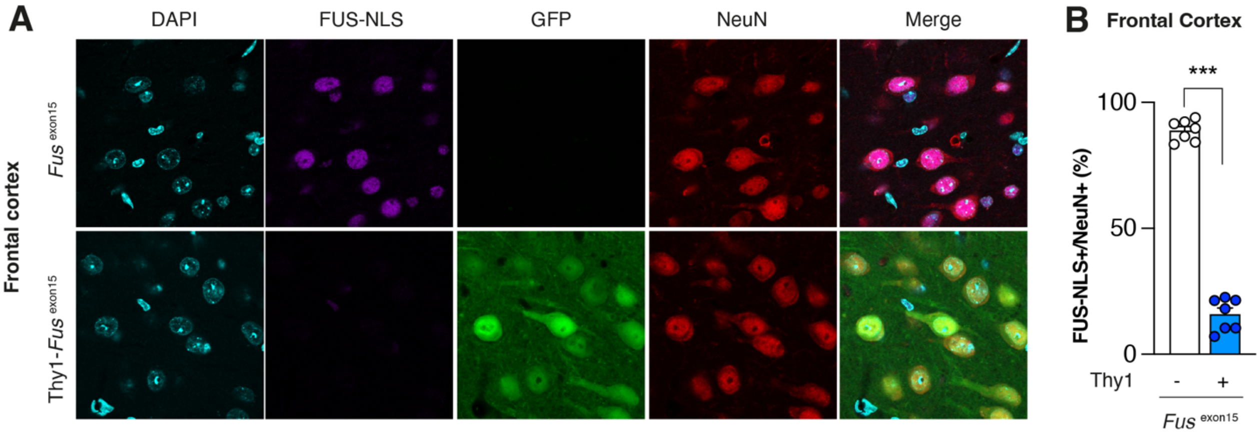


**Figure S3: complementary FUS immunoreactivity in Thy1$^{CRE\text{-}ERT2}$/ *Fus*$^{exon15}$ and control mice**
(**A**) Representative widefield images of the immunofluorescent staining of cortical neurons in the frontal cortex of 4-month-old mice. GFP and NeuN staining were used to identify recombined (GFP) neurons (NeuN). The FUS antibody used here targets the NLS-domain of FUS (deleted by the recombination). We observed a clear decrease in the number of cortical neurons positive for FUS NLS in Thy1$^{CRE\text{-}ERT2}$/ *Fus*$^{exon15}$ mice.
(**B**) Quantification of FUS-NLS+/NeuN+ cells in the frontal cortex.

Results are expressed as mean ± SEM. Statistical analyses are performed with two-tailed Mann Whitney test. ***, $p < 0.001$, n = 7 for control mice (in black), n = 7 for Thy1$^{CRE\text{-}ERT2}$/ *Fus*$^{exon15}$ (in blue).

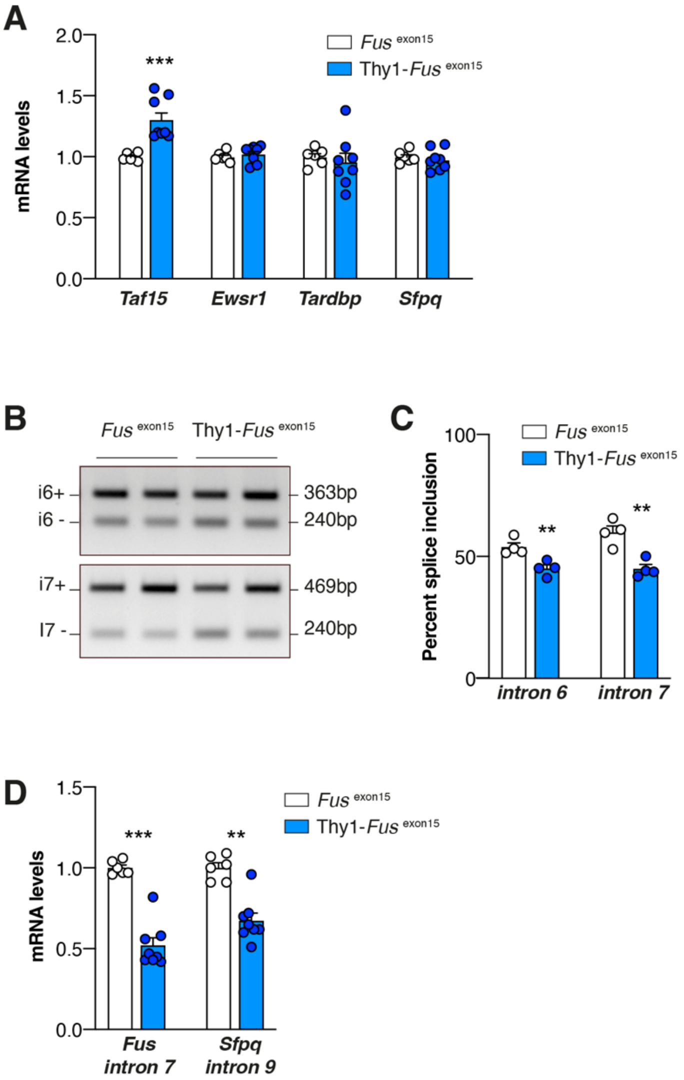


**Figure S4: complementary FUS-related molecular alterations in Thy1$^{CRE\text{-}ERT2}$/ *Fus*$^{exon15}$ and control mice**

**(A)** RT-qPCR expression of FUS-related genes. As expected, we observed upregulation of *Taf15* (***, $p<0.001$) upon *Fus* mutation while the mRNA levels of *Ewsr1, Tardbp* or *Sfpq* remained unchanged ($p>0.05$).

**(B-C)** Representative PCR (B) used to study the *Fus* autoregulatory loop and relative quantification of the percentage of splice inclusion (C). We observed a significant decrease in intron 6 and intron 7 retention in the Thy1$^{CRE\text{-}ERT2}$/ *Fus*$^{exon15}$ mice (** $p<0.01$ for both intron 6 and intron 7).

**(D)** RT-qPCR of *Fus* intron 7 and *Sfpq* intron 9 mRNAs that are two targets of FUS. We observed a significant alteration in the retention of these two *Fus*-targeted introns in the Thy1$^{CRE\text{-}ERT2}$/ *Fus*$^{exon15}$ mice ($p<0.001$ for *Fus* and $p<0.01$ for *Sfpq*).

Results are expressed as mean ± SEM. Statistical analyses are performed with two-tailed Mann-Whitney test, ***, $p < 0.001$, **, $p < 0.01$. Study of the mRNA levels (A) and FUS spliced-targeted mRNA levels (E), n = 6 for control mice (in black) and n = 8 for Thy1$^{CRE\text{-}ERT2}$/ *Fus*$^{exon15}$ (in blue). Study of Fus intron 6 / intron 7 retention (D), n = 4 for control (in black), n = 4 for Thy1$^{CRE\text{-}ERT2}$/ *Fus*$^{exon15}$ mice (in blue).

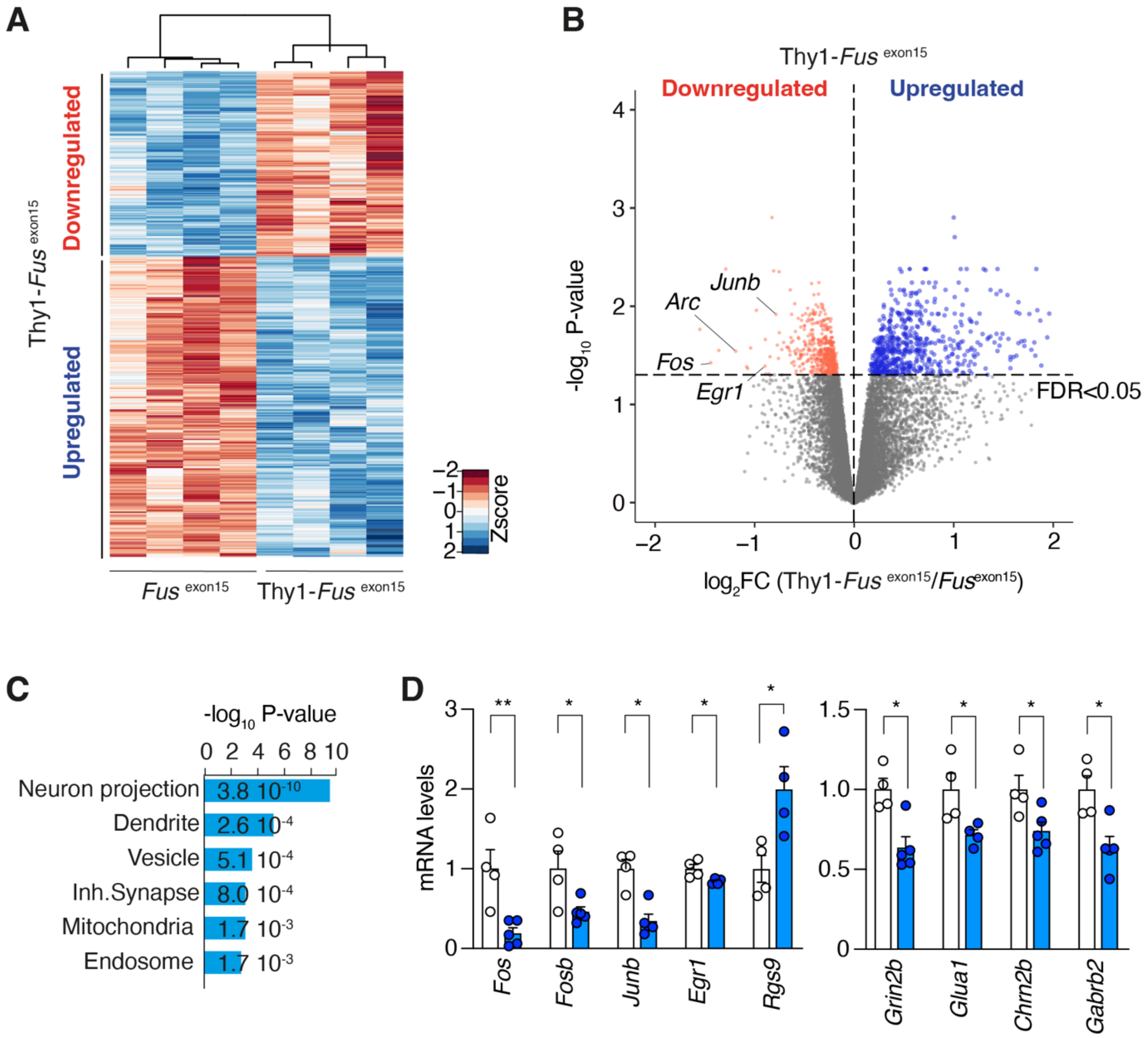


**Figure S5: RNA sequencing of Thy1$^{CRE-ERT2}$/ *Fus*$^{exon15}$ frontal cortex.**

**A-B**. Heatmaps (A) and Volcano plot (B) of differentially expressed genes in frontal cortices of Thy1$^{CRE-ERT2}$/ *Fus*$^{exon15}$ mice and their respective control littermates.
**C.** GO terms enriched in differentially expressed genes in frontal cortices of Thy1$^{CRE-ERT2}$/ *Fus*$^{exon15}$ mice.
**D.** Validation of RNAseq. We observed an important decrease of immediate early genes mRNA levels, such as *Fos* (**$p < 0.01$), *Fosb* (*$p < 0.05$) and *Junb* (*$p < 0.05$), in Thy1$^{CRE-ERT2}$/ *Fus*$^{exon15}$ mice. This was associated with alterations of pathways-regulator associated genes (*Erg1* and *Rgsp9*, *$p < 0.05$ for both) and genes involved in the neurotransmission system (*Grin2b*, *Glua1*, *Chrn2b* and *Gabrb2*, *$p < 0.05$ for all of these genes).
Results are expressed as mean ± SEM. Validation of RNAseq (D), n = 4 control mice (in black) and n = 4-5 Thy1$^{CRE-ERT2}$/*Fus*$^{exon15}$ mice (in blue). Statistical analysis: (D) Two-tailed Mann-Whitney test: ***$p < 0.001$, **$p < 0.01$, *$p < 0.05$.

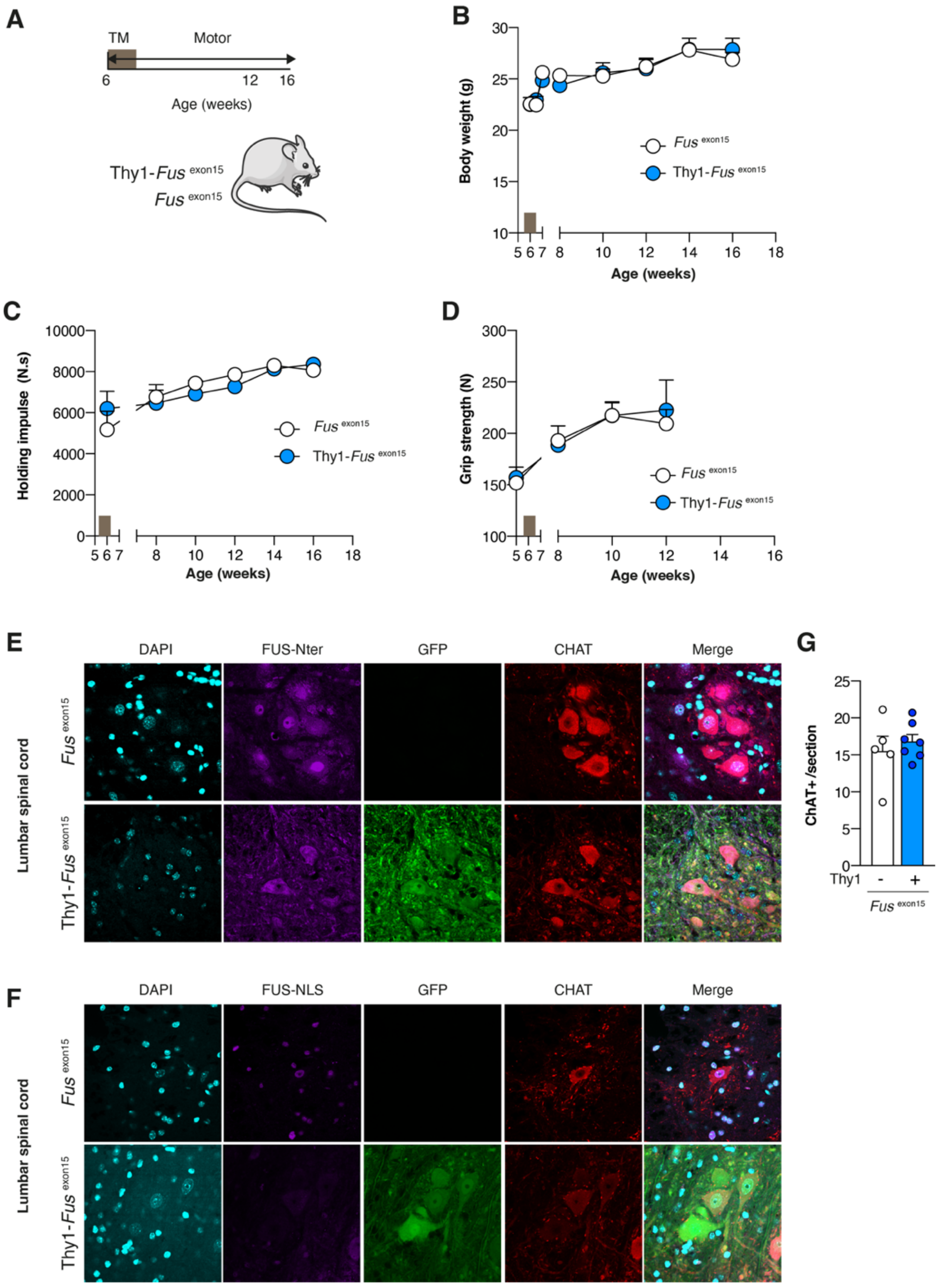


**Figure S6: Adult-onset neuronal truncation of FUS does not lead to motor deficit in mice.**

**A.** Scheme of the experimental design. The tamoxifen gavage period is indicated by the grey rectangle (TM).

**B.** Body mass was monitored daily during the induction period (left part of the graph) and then weekly during one month after the end of the induction period (right part).

**C.** Holding impulse computed from the mean hanging time. Hanging time was monitored weekly during one month after the end of the induction period as part of the inverted grid test.

**D.** Grip test was monitored daily during the induction period (left part of the graph) and then weekly for one month. (B-D) Two-way ANOVA showed no genotype effect for body weight (B, $p > 0.05$), holding impulse (C, $p > 0.05$) and grip strength (D, $p > 0.05$).

**E-F.** Representative widefield images of the immunofluorescent staining of motoneurons in the lumbar spinal cord of 5-month-old mice. GFP and ChAT staining were used to identify recombined neurons and motoneurons, respectively. We used two different stains for FUS, one recognizing the N-terminal part of FUS, i.e. the mutated and unmutated FUS (E) and the other recognizing the NLS domain of FUS (F). We observed a clear FUS delocalization in motoneurons in Thy1$^{CRE-ERT2}$/*Fus*$^{exon15}$ mice.

**G.** Number of motoneurons counted in lumbar spinal cord slices. We did not observe any decrease (two-tailed Mann-Whitney test, $p > 0.05$).

Results are expressed as mean ± SEM. Study of the weight (B) and the holding impulse (C), n = 6 control mice (in black) and n = 7 Thy1$^{CRE-ERT2}$/*Fus*$^{exon15}$ mice (in blue). Study of grip strength (D), n = 6 control mice (in black) and n = 5 Thy1$^{CRE-ERT2}$/*Fus*$^{exon15}$ mice (in blue). Study of lumbar spinal cord (G), n = 5 control mice (in black) and n = 7 Thy1$^{CRE-ERT2}$/*Fus*$^{exon15}$ mice (in blue).

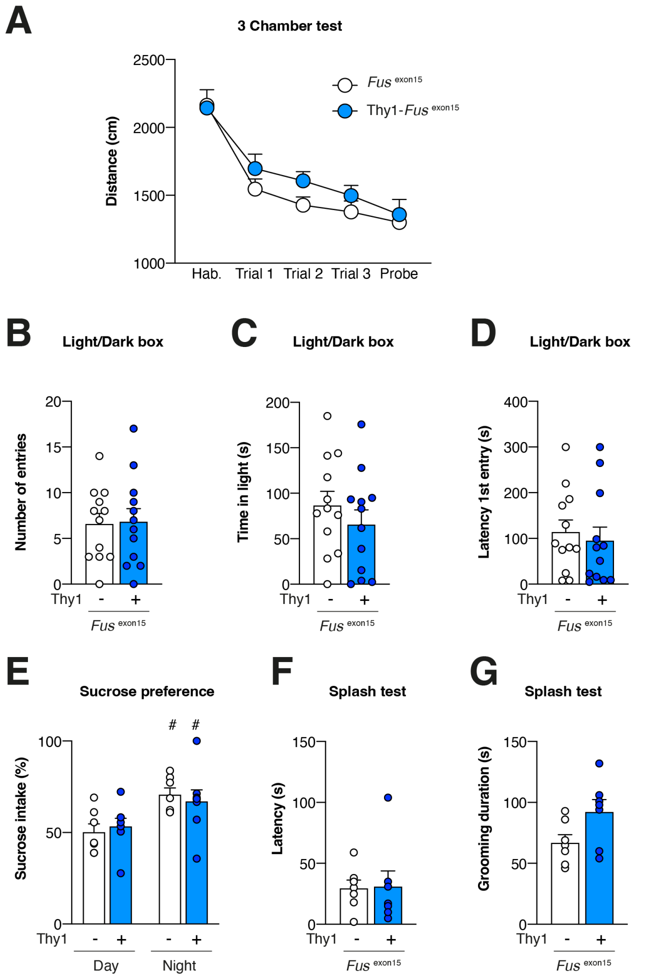


**Figure S7 Adult-onset neuronal truncation of FUS does not lead to anxiety and depressive-like behavior**

**A**. Total distance travelled during the different sessions of the 3-chamber test. There is no difference observed depending on the genotype ($p > 0.05$), but a similar decrease depending on time ($p < 0.0001$), suggesting that Thy1$^{CRE-ERT2}$/ *Fus*$^{exon15}$ mice did not suffer from major motor alterations.

**B-D**. Assessment of anxiety behavior in the Light and Dark paradigm. The number of entries in the light compartment (B), the time spent (C) and the latency to 1$^{st}$ entry (D) in the light box indicated the absence of anxiety in Thy1$^{CRE-ERT2}$/ *Fus*$^{exon15}$ ($p > 0.05$).

**E-G**. Measure of depressive-like behavior. We tested the mouse's interest in a sucrose solution (E) to measure the attractiveness of a high-palatability beverage. We found no difference depending on the genotype ($p > 0.05$) and, as expected, a higher consumption of sweetened solution during the night than during the day (##, $p < 0.01$). We finally tested the reaction of mouse when a screwing solution is applied to the fur as an indication of depression and/or anhedonia. Interestingly, the latency to start grooming is not different depending on the genotype (F, $p > 0.05$), but the total duration of grooming tends to increase in Thy1$^{CRE-ERT2}$/ *Fus*$^{exon15}$ than in control mice (G, $p = 0.057$). Results are expressed as mean ± SEM. Study of sociability with the three-chamber test, n = 6 control mice (in black) and n = 6 Thy1$^{CRE-ERT2}$/*Fus*$^{exon15}$ mice (in blue). Study of anxiety (B-D), n = 7 control mice (in black), n = 8 Thy1$^{CRE-ERT2}$/*Fus*$^{exon15}$ mice (in blue), and depressive-like behaviors (E-G), n = 7 control mice (in black), n = 7 Thy1$^{CRE-ERT2}$/ *Fus*$^{exon15}$ mice (in blue).

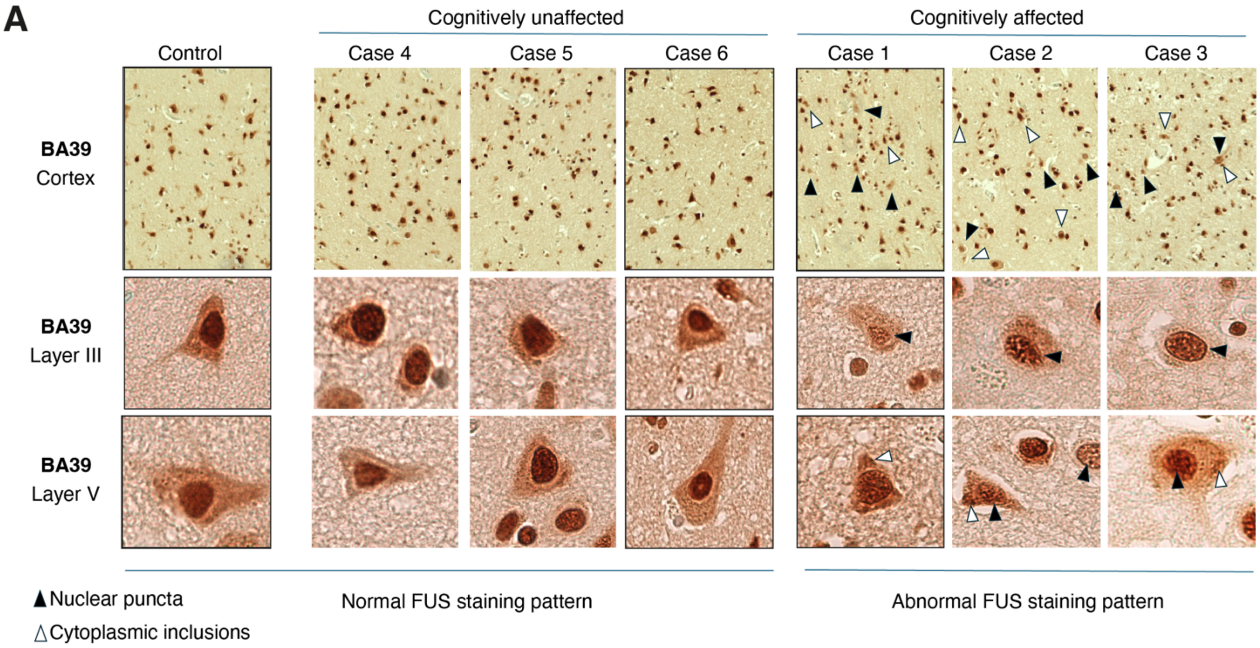


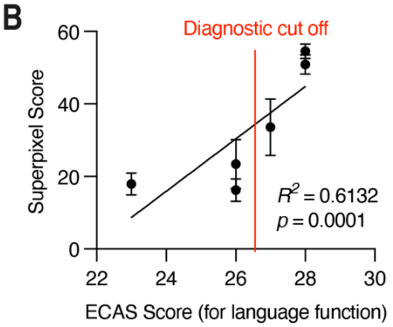


**Figure S8: additional characterization of FUS pathology in ALS and ALSci patients.**
**A.** Representative photomicrographs of *C9ORF72* human post-mortem tissue taken at 10x (top panel) and 40x magnification with optical zoom (middle and lower panels) demonstrating DAB immunohistochemical staining for FUS protein in BA39 (language brain region). Cases highlighted by a black outline are profiled in Figure A. These cases had undergone ECAS testing during life, evaluating language functions specific to this brain region, and based on clinically approved cut-offs, cases were classified as affected (ECAS score < or = 26) or unaffected (ECAS score > 26). Black arrowheads indicate nuclear pathology, and white arrowheads indicate cytoplasmic pathology within layers III and V of the cortex. Images presented in the main figures are indicated by black lines.
**B.** Correlation between the burden of pathology and ECAS score performed for that region (language function) during life. Graph shows the digital burden score of pathology performed using QuPath superpixel analysis on the y-axis and ECAS score on the x-axis. The red line indicates the cutoff for abnormality.
